# Differential impacts of contact tracing and lockdowns on outbreak size in COVID-19 model applied to China

**DOI:** 10.1101/2020.06.10.20127860

**Authors:** Cameron J. Browne, Hayriye Gulbudak, Joshua C. Macdonald

## Abstract

The COVID-19 pandemic has led to widespread attention given to the notions of “flattening the curve” during lockdowns, and successful contact tracing programs suppressing outbreaks. However a more nuanced picture of these interventions’ effects on epidemic trajectories is necessary. By mathematical modeling each as reactive quarantine measures, dependent on current infection rates, with different mechanisms of action, we analytically derive distinct nonlinear effects of these interventions on final and peak outbreak size. We simultaneously fit the model to provincial reported case and aggregated quarantined contact data from China. Lockdowns compressed the outbreak in China inversely proportional to population quarantine rates, revealing their critical dependence on timing. Contact tracing had significantly less impact on final outbreak size, but did lead to peak size reduction. Our analysis suggests that altering the cumulative cases in a rapidly spreading outbreak requires comprehensive targeted measures that decrease the reproduction number close to one, otherwise a swift lockdown may be needed.

---

The COVID-19 pandemic began in Wuhan, China, where infections grew rapidly and spread throughout the country in late December 2019 and January 2020. In order to contain the virus, drastic measures, such as travel restrictions alongside extensive lockdowns and contact tracing efforts, were implemented. The overall success of these control strategies in suppressing the outbreak in China has been recognized in several studies (1, 2). An important question is which intervention had the largest impact, or in more detail, quantifying the effect of each intervention on case reduction. The problem is relevant not only for retrospective analysis, as all countries including China face the task of controlling ongoing or possible second wave outbreaks of COVID-19, along with future emerging epidemics.

Important strategies for the fight against COVID-19 are often classified as non-pharamaceutical interventions (NPIs) because consensus vaccines or treatments may not be widely available or enough effective. The effectiveness and aims of NPIs may vary by country and type of intervention. While the goal of large-scale lockdowns and social distancing is often characterized as “flattening the curve”, whereas successful contact tracing may suppress outbreaks, a more nuanced picture of their potential impact on epidemic trajectories is necessary. A few studies have quantified impact of travel restrictions (3, 4) and lockdowns inducing large-scale changes in contact patterns or depletion of susceptible individuals (5, 6), showing the efficacy of these interventions in China. Yet, the precise qualitative and quantitative effect of brute force interventions such as lockdowns (or widespread social distancing), versus the more targeted strategy of contact tracing, on the outbreak shape is less explored.

Traditionally the influence of control strategies on outbreaks has been theoretically investigated in compartmental ordinary differential equation models of the susceptible-infected-recovered (SIR) type. Analysis yields the herd immunity (or critical vaccination) threshold for suppressing an outbreak by proportionally reducing the effective reproduction number, *ℛe*, below one, along with a nonlinear relationship between *ℛe* and final outbreak size when *ℛ*_*e*_ is above one. Furthermore, inference of parameters by fitting the model to data can help to determine the effect of interventions. However both the analytical and parameter estimation approaches are challenged by the dynamic nature of control strategies as public health authorities and individuals react to an evolving outbreak.

While the early phase of COVID-19 can be characterized by exponential growth, case saturation occurred much earlier than would be predicted by the basic SIR model due to the comprehensive control measures that have been deployed. In particular, stringent lockdown with broad (self- and contact tracing) quarantine interventions reduced the pool of susceptible individuals, effective contact rate and secondary transmissions. Several models have utilized time-dependent transmission or isolation rates to capture the dynamics (3, 7), and recent work has also considered removal of susceptible individuals at a constant rate (6). Here we develop a generalized SIR-type model incorporating a total (government mandated and individual) self-quarantine rate, along with contact tracing, both *depending on force of infection*, to fit an observed reactionary public health system and derive novel formulae for outbreak size.

To quantify the impacts of contact tracing and comprehensive social distancing (self-quarantine or lockdowns), we simultaneously utilize case and quarantined contact data from China to estimate parameters in our model. Furthermore, through computational and theoretical analysis of the model, we can explore the sensitivity of distinct epidemic measures (e.g. outbreak size, peak number of infected, timing and extent of self-quarantine) to interpretable control parameters. These investigations allow us to dissect how combinations of NPIs, such as contact tracing and lockdowns, may influence sequential outbreaks through loosening and tightening of control measures. The emergent picture is of distinct qualitative impacts of contact tracing and lockdowns on the outbreak, variable in scope and timing, and dependent on underlying disease parameters. A better understanding of these differential effects can help shape or suppress the epidemic curve of COVID-19 in a sustainable and socially acceptable manner.

## Model with Self-Quarantine and Contact Tracing

We formulate a SEIR model (Fig.1 and generalized equations are given in SI Appendix), which modifies a detailed differential equation system of contact tracing during outbreaks (8). The model variables include: susceptible (*S*), exposed (*E*) and infectious (*I*) individuals; social-distanced (or self-quarantined) susceptible (*S*_*q*_), exposed (*E*_*q*_) and infectious (*I*_*q*_) individuals; contact-traced susceptible (*S*_*c*_), exposed (*E*_*c*_), and infectious (*I*_*c*_) individuals; and the decoupled compartments of (safely) isolated reported cases (*R*) with a subset of currently quarantined contact-traced cases (*R*_*c*_). The full system of equations is given in the *SI Appendix*. Here we highlight a few key model features and parameters (Table 1). Parameters *β, β*_*c*_, and *β*_*q*_ represent transmission rates of non-quarantined, contact-traced quarantined and self-quarantined infected individuals, respectively, where *β*_*c*_ and *β*_*q*_ reflect reductions in transmission due to contact tracing and social distancing which are generally imperfect (e.g. tracing individual after they become infectious, looseness in following stay-at-home orders). A critical control parameter is the total rate of susceptible transition to (contact-traced or self-) quarantine state, *ψλS*, with dimensionless constant 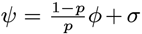, proportional to force of infection *λ* = (*βI* + *β*_*q*_ *I*_*q*_ + *β*_*c*_*I*_*c*_)*/N*, where *ϕ* is proportion of contacts traced, *p* is probability of transmission upon contact and *σ* is the self-quarantine factor. The dependence on force of infection reflects mechanism of contact tracing (8), along with the responsive nature of broader social distancing/quarantine measures to current transmission. Other important parameters include *α*_*q*_, the rate of return to susceptible from social distancing, and *ν*_*q*_ the susceptibility of self-quarantined individuals measuring the looseness of the social distancing measures.

**Table 1.** Key model parameters and quantities.

|  |  |
| --- | --- |
| $\lambda = \frac{\beta I + \beta_q I_q + \beta_c I_c}{N}$ | force of infection of non-quarantined, self-quarantined & contact-traced infected. |
| $\phi$ | Proportion of contacts traced & quarantined |
| $\sigma$ | susceptible self-quarantine factor |
| $p$ | probability of transmission upon contact |
| $\psi = \sigma + \frac{1-p}{p}\phi$ | total susceptible quarantine factor (proportionality constant relative to force of infection) |
| $\nu_q$ | Relative susceptibility of self-quarantined |
| $\alpha_q, \alpha_c$ | Exit rate from self-, contact-traced quarantine |
| $\tau$ | time to infectious (incubation period) |
| $T, T_q, T_c$ | infectious period (time to isolation) of non-quarantined, self-quarantined, contact-traced |
| $\mathcal{R}_{0,b}, \mathcal{R}_0, \mathcal{R}_e(t)$ | reproduction numbers (baseline, with contact tracing, effective). |

A simplified version of the model assuming *perfect indefinite* quarantine and *perfect* contact tracing is given by the following system:

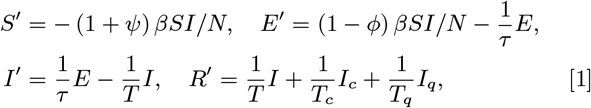

where *R* (fit to data) is decoupled, and the additional decoupled compartments of self-quarantined and contact-traced (fit to data) are detailed in the SI. Note that for superior analytical results and model identifiability, for simplicity we assume that the infectious and exposed period (*T* and *τ*) remain constant. This can be interpreted as averaging an evolving serial interval (e.g. due to improving non-pharmaceutical interventions (9)) over the outbreak timeframe (see also *SI Appendix* for alternative non-constant serial interval model and fitting).

## Reproduction Number and Outbreak Size

The *reproduction numbers, ℛ*_0,*b*_ and *ℛ*_0_, (baseline and after contact tracing) of the “perfect quarantine” model Eq. (1) are:

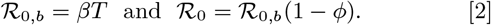

We also consider (time-dependent) *effective reproduction number ℛ*_*e*_ = (*S*(*t*)*/N*) *ℛ*_0_, calculated continuously during the outbreak. The *R*_*e*_ for the full model (Fig. 1) depends upon the quarantined populations *S*_*q*_ (*t*) and *S*_*c*_(*t*), along with their imperfect susceptibility and transmission parameters (*ν*_*q*_, *ν*_*c*_ and *β*_*q*_, *β*_*c*_) and is formulated in *SI Appendix*.

**Fig. 1.**
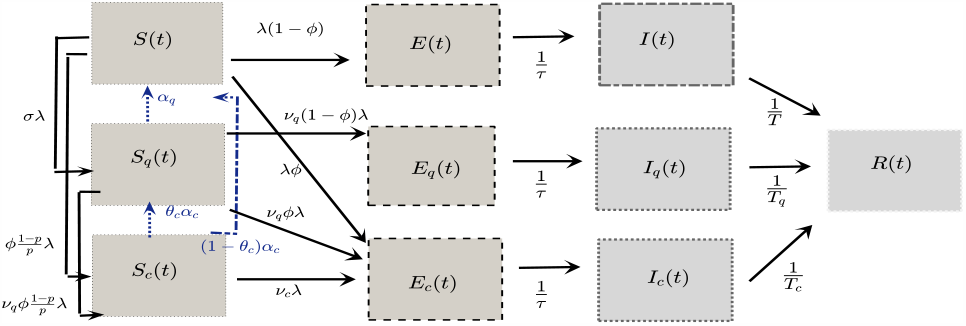
Full model of COVID-19 with reactive contact tracing and self-quarantine. We extend the basic SEIR model describing Susceptible (*S*), pre-infectious Exposed (*E*), Infectious (*I*), and Reported case (*R*) compartments to also include contact-traced (*S*_*c*_, *E*_*c*_, *I*_*c*_) and (lockdown or social distancing following) self-quarantined (*S*_*q*_, *E*_*q*_, *I*_*q*_) individuals. See Table 1 and *SI Appendix* for model descriptions, and Eq. (1) for simplified “perfect quarantine” system.

Next we present novel analytical relations between *ℛ*_0_, self-quarantine rate and final or peak outbreak size. We assume an indefinite quarantine period (*α*_*q*_ = 0), as in Eq. (1). In this way, the outbreak size can represent magnitude of first, second or subsequent waves dependent on contact tracing and self-quarantine parameters. Define the final proportion of susceptible individuals 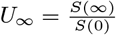, the *final (cumulative) epidemic size* 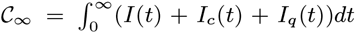, and peak non-quarantined infected (and exposed) individuals *𝒴*_*peak*_:= max_*t>*0_ *(E(t) + I(t*)). Under simplifying conditions, we derive the final size of *U*_*∞*_ and *𝒞*_*∞*_ dependent on the (assumed) uniform susceptibility of self-quarantined and contact-traced individuals, *ν* (detailed in *SI Appendix*). In the best case scenario where self-quarantine or contact tracing perfectly prevents susceptible infection (*ν* = 0), we obtain the exact formulae:

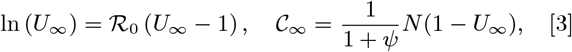

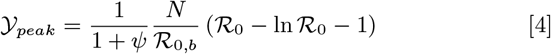

where *ψ* depends on self-quarantine factor (*σ*) and *ℛ*_0_ depends on contact tracing proportion (*ϕ*). Note that each formula can account for arbitrary initial conditions in order to quantify how *ℛ*_*e*_ and quarantine measures affect outbreak or peak size beginning at any stage (see *SI Appendix* for formal derivations).

In Eq. (3), note the classical relation between final or peak outbreak size and *ℛ*_0_ when *ψ* = 0. Self-quarantine (*σ*) is expected to have almost all weight in the total susceptible quarantine factor (*ψ* = *σ* + *ϕ*(1 − *p*)*/p*) because the goal of lockdowns, as opposed to contact tracing (*ϕ*), is to shield large segments of susceptible population. Thus both final and peak outbreak size, *𝒴*_*peak*_ and 𝒞_∞_, have a simple inverse proportionality relationship with *σ*. In contrast, contact tracing, primarily acting through the proportional reduction in baseline reproduction number (*ℛ*_0_ = (1−*ϕ*) *ℛ*_0,*b*_), has distinct influences on peak and final size. We calculate relative sensitivities of Eq. (3)-Eq. (4) to *ψ* and *ϕ*,

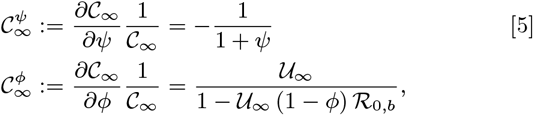

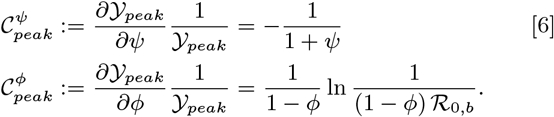

Observe that as *ℛ*_0_ increases, contact tracing has more influence on peak size, but less impact on final size, where relative rate of change of *𝒞*_∞_ with respect to *ϕ* goes to zero as *ℛ*_0_ becomes large.

Although there are inherent issues with measuring rate of new incidence due to the delay between transmission and case reporting, the inverse proportions dependent on susceptible quarantine factor (*ψ*) can be used as a simple retrospective or implementation guide for lockdown efficacy. For instance, given that *ψ* is relative to force of infection, to reduce the outbreak size by 90% (compared to no lockdown case *ψ ≈*0), the authorities would need to have implemented strict quarantine at approximately 9 times the force of infection. Note that *ℛ*_0,*b*_ is the ratio of force of infection 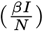 and reported prevalence rate 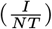. Thus, as a rule of thumb, the quarantine rate (or magnitude in a defined time period) must be 9 × *ℛ*_0,*b*_ times the rate (or amount) of incoming reported cases to reduce the outbreak size by 90%. In practice, officials may look at aggregated cell phone or mobility data to assess population quarantine behavior and compare with case reports. Quarantine rate may need to far exceed the pace of new infections, as shown below, very large values of *ψ* were instrumental for China rapidly curbing their epidemic.

## Data Fitting & Distinct Quarantine Efficacies in China

In order to quantify relative efficacies of contact tracing and self-quarantine (lockdown) interventions during the COVID-19 outbreak in China, we simultaneously fit our models to daily reported case data and (daily number of) quarantined contacts from daily reports by NHC (10). The outbreak was not localized to a single population during the timeframe of our study, Jan. 21-Mar. 19 2020, although the province of Hubei had a large share of the total cases. For each province in China we fit model Eq. (1) to daily case incidence, along with an inferred number of quarantined contacts, weighting the available aggregated quarantine contact data for all of China by province reported case load distributed by an assumed exponential quarantine duration of mean 14 *days* (see *SI Appendix*). To increase model identifiability, we reduced the number of parameters by fixing the incubation period (time to infectiousness, *τ* = 3 days (11)), infectious period (time to isolation, *T* = 4.64 days (12)), and probability of transmission given contact, *p* = .06, in line with other studies (12, 13).

The province model fits (see *SI* table S4, figures S8,S9) can largely be summarized by separating daily incidence into Hubei and China less Hubei. Thus we refined our analysis by fitting their two corresponding daily reported case datasets and the sum of their respective quarantine contact compartments to the aggregated China daily quarantine contacts (Fig. 2). The estimated basic reproduction numbers (with contact tracing) are *ℛ*_0_ = 4.07 (*CI* 3.54 −4.12) and *ℛ*_0_ = 2.47 (*CI* 2.16 − 2.77) for Hubei and China less Hubei, respectively. Complete parameter values and uncertainty analysis are presented in *SI Appendix*. We also calculate time-dependent *ℛ*_*e*_ by directly utilizing the daily case data and estimates of the serial interval (generation time) distribution (8, 12, 14), alongside *ℛ*_*e*_ obtained from our estimated parameters from model fitting. Although China less Hubei had larger contact tracing level than Hubei (*ϕ* = 0.59 versus *ϕ* = 0.32), possibly due to smaller caseloads, it also had significantly higher estimates of self-quarantine rate (*σ* = 1.14 10^5^ versus *σ* = 1.26 10^3^) likely attributed to the speed advantage in reacting to the initial outbreak in Hubei in addition to their local cases. The results indicate strict population-wide lockdowns were the main quarantine measure (as opposed to contact tracing) which rapidly contained the outbreak in China by rapidly decreasing *ℛ*_*e*_ (see Figs. 2(a),2(b)).

**Fig. 2.**
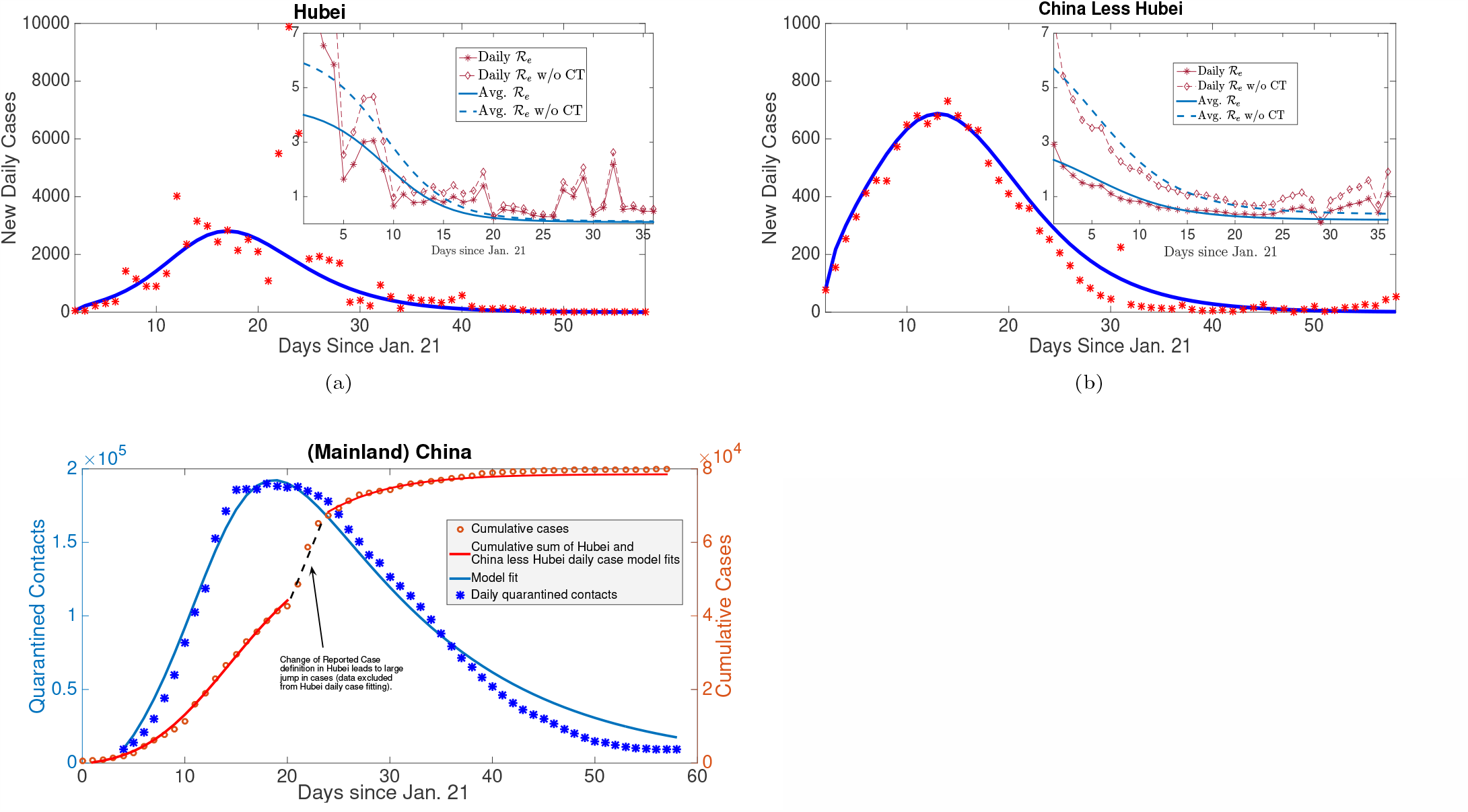
Model recapitulates case and contact tracing data, estimates large rate of self-quarantine (via lockdown) rapidly contains outbreak. (a),(b) Daily reported cases (incidence) of Hubei and China less Hubei fit to model Eq. (1). Also, average and daily impact of contact tracing on *ℛ*_*e*_ (inserted figures) from estimated parameters and statistical generation time distribution approach. (c) Daily number of quarantined contacts in mainland China simultaneously fit with daily incidences of Hubei and China less Hubei (in (a),(b)) shown alongside inferred cumulative cases from sum of incidence fits and data. By quantifying relative magnitudes of contacts traced and incidence, we can determine efficacy of contact tracing versus broader lockdowns in our distinct quarantine model. The reduction in *ℛ*_*e*_ with or without contact tracing due to self-quarantine/lockdowns was the major factor to rapidly contain the outbreak in China.

Next, we observe an inverse proportionality relationship between province outbreak size and corresponding estimated self-quarantine factor (*σ*), supporting theoretical predictions of our final size formula Eq. (3) (Fig. 3). Indeed, the 30 provincial cumulative case counts over the study period are inversely proportional to fitted *σ* values, whereas contact tracing (*ϕ*) did not have a significant correlation with province outbreak size. This inverse relationship was also true of province peak size versus self-quarantine factor, but contact tracing had a larger impact here than for the case of cumulative cases, as illustrated by distinctly flatter peak size sensitivity curves (with respect to varying *σ*) for provinces with larger proportion of contacts traced, *ϕ*. The small effect of contact tracing on final size, but larger influence on peak size, concurs with our derived sensitivity analysis when *ℛ*_0_ is relatively large. To further corroborate the heterogeneous impacts of contact tracing, we calculate outbreak sizes if *ϕ* = 0, finding peak infection reductions of 28% in Hubei versus 45% in China less Hubei. However, the decrease in final outbreak size due to contact tracing was only 1.6% (Hubei) and 5.6% (China less Hubei), respectively.

**Fig. 3.**
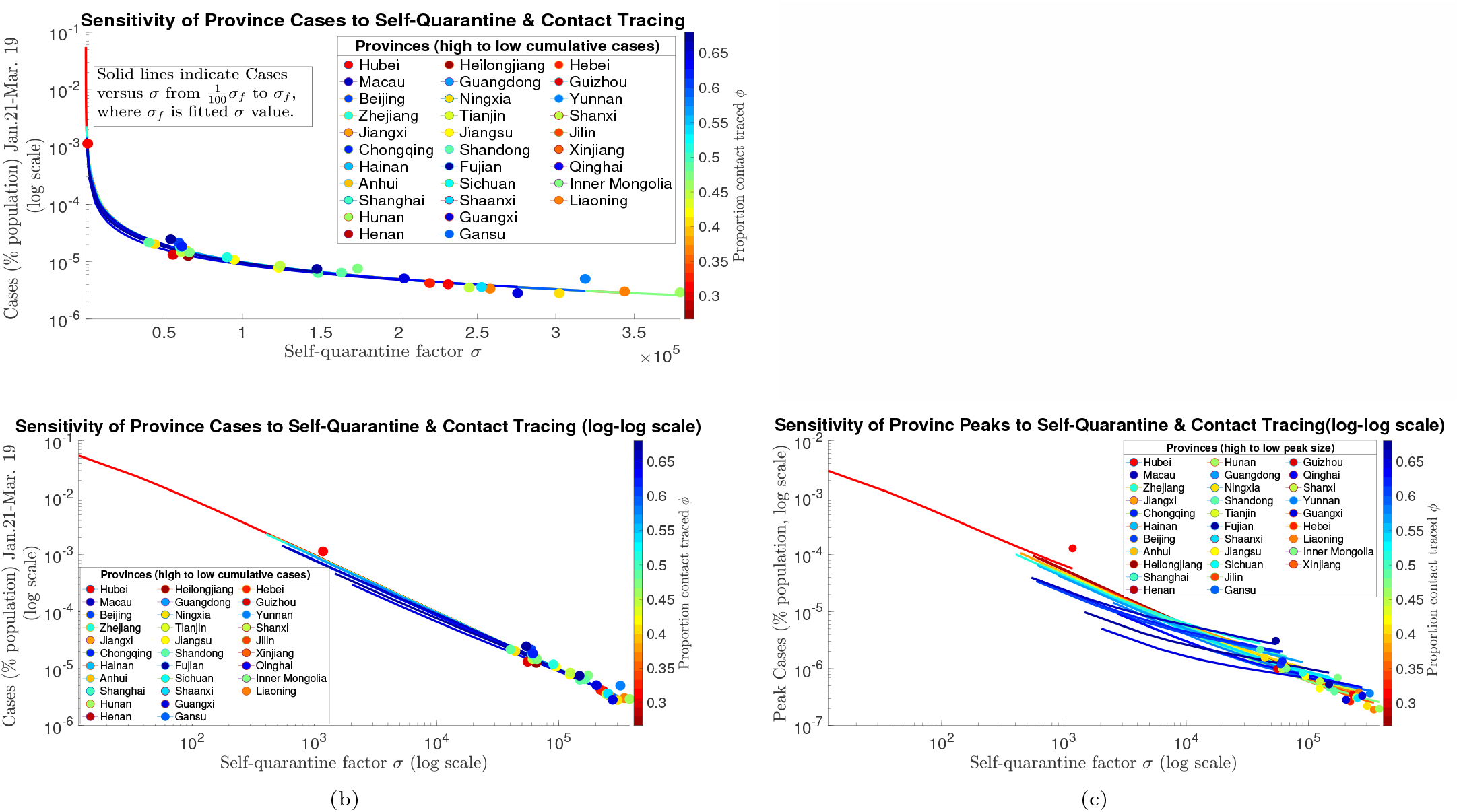
Model fitting shows that COVID-19 outbreaks in Chinese provinces mimic derived inverse proportionality relationship with self-quarantine rate and limited differential impacts of contact tracing. (a) Data points represent cumulative reported cases during Jan. 21-Mar. 19 of 30 provinces in China plotted at estimated self-quarantine factor (*σ*) obtained from provincial daily incidence data and inferred daily number of quarantined contacts (weighted from aggregated China data) simultaneously fit to model Eq. (1). The solid lines represent calculated cumulative cases from final size formula Eq. (3) while varying *σ* from 1% to 100% (*fitted value*) of the estimated *σ* (*σ*_*f*_) and fixing other parameter estimates for each province. All province output is colored according to a colormap from minimum to maximum estimated contact tracing parameter *ϕ*. (b) The same output as (a) displayed on log-log scale. (c) Analogous graph as (b) with respect to peak size for each province instead of cumulative cases (again displayed on log-log scale to aid viewing distinct province curves). Note the distinctively flatter peak size curves (with respect to varying *σ*) of provinces with high estimated contact tracing (*ϕ*) demonstrating the larger influence of contact tracing in keeping peak size small even as the lockdown scale (*σ*) decreases.

A change in case counting procedures around Feb. 12 in Hubei province prevented us from reliably fitting daily case counts in this province from Feb. 11-Feb. 13, and thus we also performed our fitting procedure on cumulative cases, rather than daily incidence, in order to obtain an overall picture of the total outbreak in China. The initial amount of infected individuals, *I*_0_, was estimated at 778 on January 21, 2020, when the dataset begins around the time when major lockdowns began (e.g. a *cordon sanitaire* implemented in Hubei province on Jan. 23). However our force-of-infection (*λ*) dependent rate formulation of mass self-quarantine (*σλ*) actually allows for a similar epidemic trajectory when instead initiating the model a month earlier with one infected individual (see *SI Appendix* Fig. S1). Furthermore, we verified robustness of our parameter estimations and results by fitting the full model ((1) in *SI Appendix* and Fig.1) simultaneously to cumulative reported cases and daily quarantined contacts in all China, along with exploring alternative formulations of self-quarantine rate (e.g using mobility data) and different quarantine/infection residence time distributions, and decreasing infectious periods due to lockdowns (see *SI Appendix*). Despite the various modeling assumptions and the inconsistencies in the reported case data, our results on how contact tracing and self-quarantine/lockdown measures affected outbreak and peak size were robust (see *SI Appendix* for full details).

Sensitivity analysis on the main control parameters for the all China fit (Fig. 4) reveals minimal contact tracing level impact on final outbreak size, yet larger effect on reducing peak infection levels (Fig. 4(g)), concurring with our analytical calculations (Eq. (3),Eq. (4), Eq. (5), Eq. (6)) and results when fitting province daily incidence. In general, we observe that the time to peak increases with *ϕ*, reflecting the curve flattening, however this time period eventually decreases for sufficiently large values of *ϕ* as contact tracing effectively suppresses the outbreak (Fig. S7). In addition, with sufficiently large contact tracing coverage, outbreak size can be significantly reduced when there is less stringent lockdown (less total quarantined and more time to enact quarantine). Yet even in this case, some level of broader social distancing measures is almost certainly needed in combination with contact tracing.

**Fig. 4.**
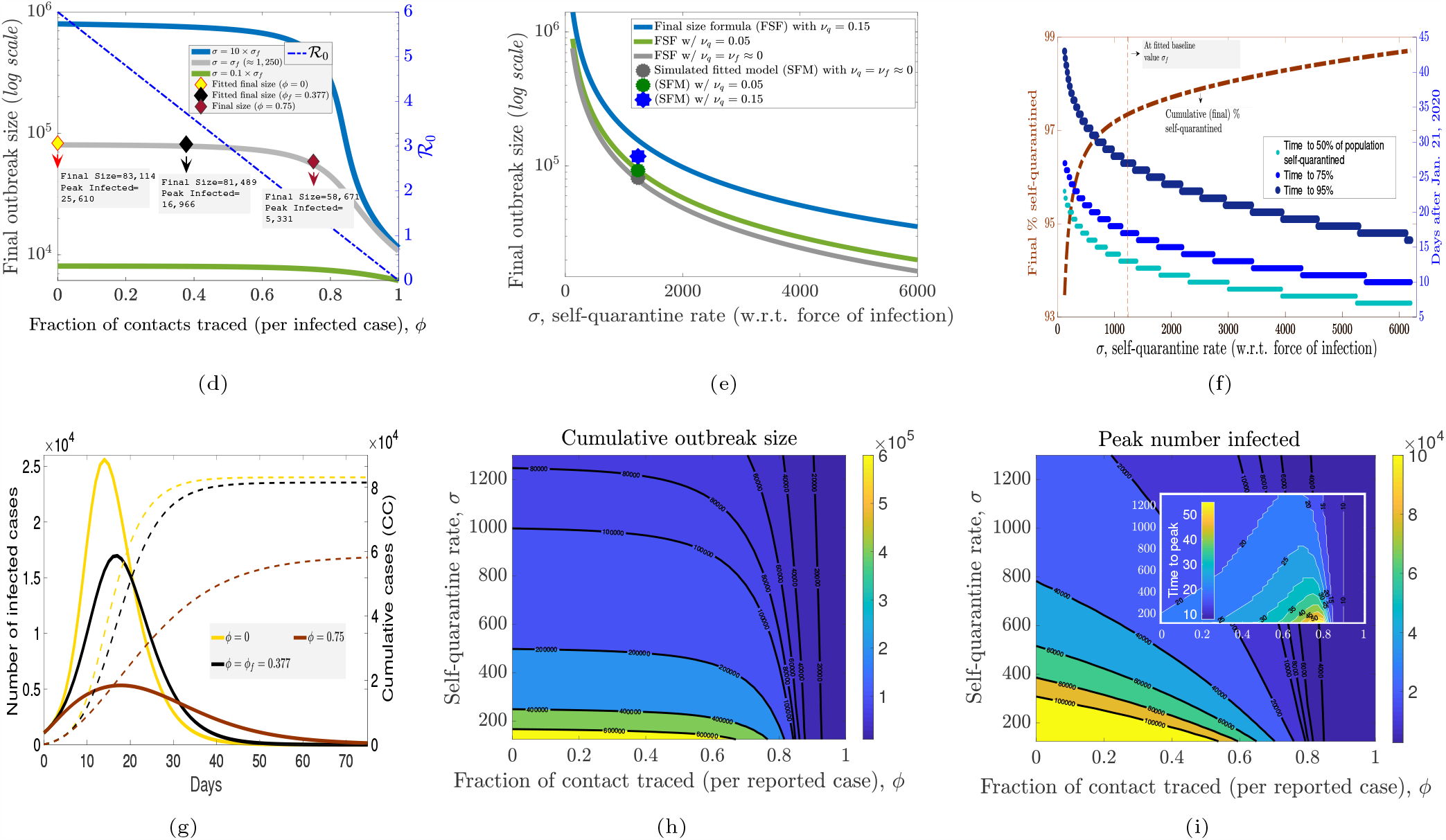
Sensitivity analysis of contact tracing and self-quarantine. All together the figures illustrate how contact tracing (*ϕ*) has less impact on outbreak size (*C*_*∞*_) than self-quarantine (*σ*) due to underlying concavity of their nonlinear relationships derived in final size formula Eq. (3). Fitted parameter values for total cumulative cases in China show that rapidly implemented lockdowns contained the outbreak, where a delay of 2 weeks in 95% of susceptible population being self-quarantined would result in a 10-fold increase in cumulative cases. Although contact tracing had little effect (2%) on final outbreak size, it “flattened the curve”, reducing peak infected by 34%. (a) Contact tracing (CT) proportion *ϕ* versus outbreak size *C*_*∞*_ (nonlinear relationship) and reproduction number *ℛ*_0_ (linear relationship) for 3 levels of self-quarantine (SQ) factor *σ* differing by order of magnitude of 10. (b) *C*_*∞*_ as a (nonlinear) function of *σ* for 3 levels of SQ susceptibility reduction *ν*_*q*_. (c) Total (final) self-quarantined individuals, along with time until 50%, 75%, 95% of population is self-quarantined as SQ factor *σ* varies. (d) Epidemic curve trajectories for 3 levels of CT proportion *ϕ* marked in (a). (e,f) Contour maps of outbreak size and peak infected (with time to peak inserted) depending on CT proportion *ϕ* and SQ rate *σ*.

As predicted by our derived inverse proportionality relationship Eq. (3), there are escalating costs as *σ* decreases, i.e. as self-quarantine action lessens relative to ongoing infection rate (Fig 4(e)). For example, if the estimated time for 50% of initial susceptible population of China to be self-quarantined (*∼*2 weeks from Jan. 21) had been delayed by just one week, then the total number of cases would be approximately 10 times larger (Fig. 4(f)). In the full model, the additional parameters *ν*_*q*_, *β*_*q*_ (measuring looseness of the lockdown) also impacts the outbreak size (Fig. 4(e)). Going from *ν*_*q*_, *β*_*q*_ */β ≈* 0 (as predicted for China) to *ν*_*q*_ = *β*_*q*_ */β* = 0.05 to *ν*_*q*_ = *β*_*q*_ */β* = 0.15, the outbreak size would increase by a factor of 1.14 and 1.45, respectively. In addition, the estimated exit rate from self-quarantine (*α*_*q*_) for China was estimated to be very small, emphasizing the strictness of the lockdown.

## Quarantine Interventions for COVID-19 2nd Wave

A major question is how lockdown measures should be loosened after some level of controling COVID-19, while optimally responding to any subsequent outbreaks induced by the relaxations. Here we analyze how the scale and rate of different reactive contact-based interventions affect 2nd wave outbreaks under two different scenarios of loosening, namely *Instantaneous Return of Several Sectors* (IRSS) or via *Gradual Return of Self-Quarantined* (GRSQ). The goal is to attain qualitative insights on strategies for shrinking, flattening or delaying (not necessarily independent phenomena (15)) subsequent outbreaks. By varying self-quarantine factor *σ*, contact tracing probability *ϕ* and looseness of social distancing *ν* under the distinct relaxation policies in our model parameterized to data from China, we observe potential consequences of different measures.

Simulating the instantaneous return of 80% of self-quarantined individuals (IRSS strategy), with no change in parameters (and crucially the same “reactive lockdown” factor *σ*) we observe that the cumulative number of infected cases for the 2nd wave (outbreak size) and peak infected was 75% and 58%, respectively, of the 1st wave. Furthermore, a similar number of individuals as during the first wave lockdown re-enter self-quarantine about 6 weeks after relaxation (see Fig.5(a)). When the contact tracing efforts are enhanced after lockdown (to *ϕ* = .65), outbreak size and peak infected are 54% and 16%, respectively, of the 1st wave, and the curve is flattened, i.e. the peak outbreak size shrunk and the time to peak outbreak size increased. When contact tracing is doubled to *ϕ* = 0.75, or if other non-quarantine measures (e.g. face mask usage) reduce*ℛ*_0,*b*_ to 3 and *ϕ* = 0.5, the 2nd wave outbreak size and peak infected are 25% and 3%, respectively, of the 1st wave. In addition, the number of individuals re-entering self-quarantine was reduced, revealing that contact tracing can be an effective tool for managing the epidemic with a less stringent lockdown.

In the case of GRSQ strategy, after containing initial outbreak with lockdown, we increase the return to “normalcy” rate to *α* = 0.01, where half the social-distanced return to normalcy in the approximate half-life time given by *t*_1*/*2_ = ln 2*/α* = 72 *days*. Assuming other parameters remain constant (including the reactive SQ factor *σ*) the second peak, emerging with a 100 day delay, reduced to 42% of the first wave, however the number of infected individuals settle into a rather large quasi-equilibrium resulting in more cumulative cases (see Fig.5(d)). Here there is a balance of force of infection induced self-quarantine (*σλ*) and reversion of individuals to their normal contact behavior (*α*), leading to an insufficient amount of population social distancing for reducing cases below a certain level. On the other hand, after loosening the lockdown, when the contact tracing efforts are enhanced or doubled, the peak size significantly diminished (27% or 0.3% of 1st wave), along with the number of self-quarantined. Importantly, for about 6 months (or the whole year in the case of doubling *ϕ*), the number of infected cases stayed significantly low. This suggests that gradual release of self-quarantined individuals with increasing contact tracing efforts can be used as a strategy to gain time until vaccination, while reinstating societal interactions in a carefully measured stepwise fashion.

Responsive re-implementation of lockdown (or social distancing) measures is crucial for reducing any second wave outbreak. Reduction in SQ factor *σ* by 1*/*2 (or 1*/*4), as predicted simply by the inverse proportionality in the derived final size formula Eq. (3), results in twice (or four times) more cumulative cases for the 2nd wave, and the simulations show the same relations between peak size (see Fig.5(b)). Although the number of self-quarantined individuals eventually become the same with the different SQ rates, the delay in implementing large-scale self-quarantine (in response to incidence) makes significant differences in the final (and peak) outbreak size. For the simulations presented in Fig. 5(b), a delay of just 9 days from the baseline parameter case results in twice as many infections, and a delay of 18 days induces four times the infected individuals. Compared to instantaneous release, the increased quarantine exit rate (*α*) under gradual return resulted in larger (but delayed) peak and total outbreak size inversely proportional to declines in SQ factor *σ* (see Fig.5(e)). Finally, varying the looseness of the quarantine (measured by uniform susceptibility and infectivity values *ν*_*q*_ = *β*_*q*_ */β*) from perfect quarantine to 25% (or to 50%) looseness, leads to approximately 1.3 times (or to 2 times) more total and peak infections during the outbreak. Different from the rate of SQ, the proportionality relations are nonlinear, thus a slight looseness in social distancing can still offer an effective intervention, but the cases will increase at a growing rate as the measures become less strict (see Fig.5(c),5(f)).

**Fig. 5.**
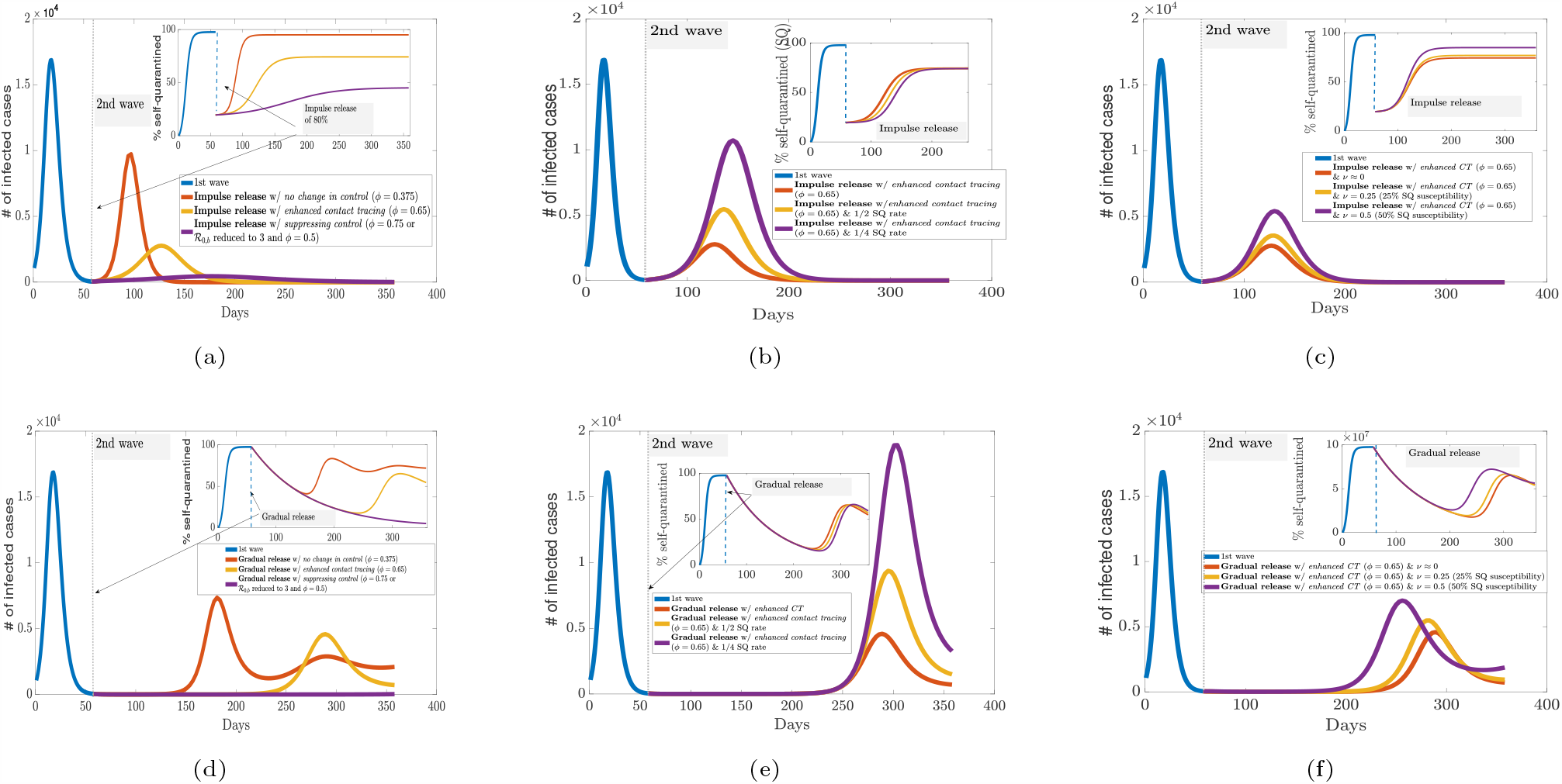
Combined impact of contact tracing and responsive social distancing (self-quarantine) measures under distinct re-opening strategies. Model simulations predict 2nd wave outbreak shape after instantaneous re-normalization of several sectors (IRSS) by releasing 80% of self-quarantined (a,b,c) versus gradual return of self-quarantined (GRSQ) to normalcy (d,e,f). Increased contact tracing levels (*ϕ*) can flatten or suppress (when highly effective) subsequent outbreak in (a,d). Social distancing measures responsive to new incidence shrink the outbreak size dependent on rate (*σ*) and looseness (*ν*_*q*_) of self-quarantine in (b,e) and (c,f), respectively. In addition to the differential effects on daily infected counts of contact tracing and responsive self-quarantine (flattening versus crushing), observe the much longer delay in 2nd wave under the GRSQ re-opening policy, which can buy time for effective treatments or vaccines.

## Discussion

In this study, we compare how two distinct types of contact-based interventions, namely contact tracing and large-scale lockdowns/self-quarantine (or social distancing), impact single or sequential COVID-19 outbreaks. We find that contact tracing generally is less effective in decreasing outbreak size for rapidly spreading pathogens (high baseline reproduction number *ℛ*_0,*b*_), unless the tracing is very efficient and large scale, or combined with other measures to force *ℛ*_0_*≈*1. On the other hand, widespread lockdowns/social distancing interventions can lower outbreak size inversely proportional to an increase in the rate of self-quarantine. Our analysis indicates that China benefited from the heavy influence of lockdowns by rapidly containing the quickly growing COVID-19 cases, and, despite massive efforts, contact tracing was less influential in bringing down the epidemic.

Despite the difference in the targeted nature of contact tracing versus the more indiscriminate lockdown measures, we contend there is a similar reactive quality to both control strategies. Contact tracing reacts to reported cases by tracking and (to varying degrees) quarantining individuals whom have been contacted. Mass social distancing or self-quarantine reflects a natural response by both governments and individuals that intensifies as cases build, a phenomenon that has been labeled as “exponential whiplash” (16). These features motivate us to construct a COVID-19 model with both contact tracing (mechanistically) and self-quarantine (phenomenologically) dependent on force of infection. In contrast to another model that assumes a linear rate of self-quarantine (6), the nonlinear social distancing rate captures a contagion-like behavioral response to infected cases, and allows us to derive novel formulae for final outbreak size. Furthermore the model provides a good simultaneous fit to both daily reported case and quarantined contact data from China.

An important distinction between contact tracing and lock-downs is their mode of action, namely preventing onward secondary infections by early tracking of likely infected cases in the former and large-scale depletion (or shielding) of susceptible individuals for the latter. This contrast determines how they affect the major epidemiological quantities of reproduction number and outbreak size in our “transmission-reactive” formulation. In particular, contact tracing proportionally reduces *ℛ*_0_, akin to vaccination, leading to a nonlinear relationship with final outbreak size, which decreases substantially only as *ℛ*_0_ approaches one. The responsive self-quarantine factor does not affect *ℛ*_0_, and we derive a simple inverse proportionality with outbreak size. This can be translated to a time of action for quarantine measures, analytically demonstrating the escalating impacts of delaying implementation of responsive lockdowns beyond a critical time period, which has been observed in other studies via simulation (17, 18). Even though similar levels of self-quarantine would eventually be reached in our model as incidence grows, the cost of delays can result in a large excess of cases.

Our main result on differential impacts of lockdowns and contact tracing also implicates distinct metrics for officials to monitor when enacting each intervention. For reactive lockdowns, the key measurement is rate of population-wide quarantine compared to incidence or incoming reported cases. For contact tracing, *ℛ*_*e*_ may be the important quantity because the final outbreak size is only substantially altered as *ℛ*_*e*_ becomes closer to 1. Although we find that the extensive lockdowns were a much larger factor in controlling COVID-19 *outbreak size* in China, our sensitivity analysis shows that contact tracing did dampen and delay *peak number of infected* despite its more limited impact on the cumulative count. In this way, contact tracing flattened the incidence curve, easing the strain on limited hospital resources. A combination of expeditiously enacted contact-based interventions may be the best strategy, where effective contact tracing and responsive social distancing measures can synergistically and efficiently suppress an outbreak. However COVID-19 has proved to be a particular challenge and large-scale lockdowns have been a needed antidote for controlling outbreaks in several countries. Drastic self-quarantine orders can also reduce case numbers to manageable levels that allow for effective contact tracing after easing restrictions.

The capacity to respond to the continuing threat of COVID-19 will be vital for minimization of sequential epidemic waves. We investigated control measures under an instantaneous normalization of contact for a large portion (or several sectors) of the population versus a more gradual release of self-quarantined individuals back into social interactions. Our results show that increased contact tracing efforts can alter the second outbreak shape, either reducing and spreading out the number of infected or completely suppressing cases for highly efficient tracing. Social distancing or lockdown measures responsive to incidence can effectively compress the second peaks, with the timing being critical again. Either measure will depend upon sufficient case detection and reporting, highlighting the importance of testing. Furthermore, indefinite or reoccurring strict lockdowns are likely to impart too high of an economic cost, and our model shows that looser restrictions and contact tracing can still control a second wave. Additionally, the strategy of gradual release of quarantined sectors can substantially delay the second wave, possibly buying time for effective treatments or vaccines to be developed.

A major part of this work was to incorporate data on the quarantined contacts compiled for all of China. Obtaining provincial quarantine records or more detailed contact tracing data quantifying the proportion of reported cases whom were traced can allow for superior accuracy in estimating efficacy of contact tracing. A recent study of detailed contact tracing records in Hunan province of China (13) did come to a similar conclusion as our work that lockdowns had larger impact, and contact tracing alone is not likely to be sufficient for controlling COVID-19. The results here explain the differential effects that contact tracing and reactive population-wide self-quarantine have on reducing cumulative cases in a rapid spreading outbreak. This knowledge and further investigation may offer insights for the public health response to COVID-19.

## Data Availability

Source code data have been deposited in GitHub (https://github.com/jcmacdonald-codesData?tab=projects).

## Supporting Information (SI)

SI Appendix

Figs. S1 to S25

Tables S1 to S5

## ACKNOWLEDGMENTS

CJB, HG, and JCM are supported by a U.S. National Science Foundation RAPID grant (DMS-2028728). HG was also supported by an NSF grant (DMS-1951759) and a grant from the Simons Foundation/SFARI(638193). CJB is partially supported by an NSF grant (DMS-1815095). We thank Fadoua Yahia for insightful discussions on deriving outbreak size formulae and sensitivities.

