## Supplementary material for "Differential impacts of contact tracing and lockdowns on outbreak size in COVID-19 model applied to China": SI Appendix

### 1. Methods

**A. Model description.** Here we formulate our full generalized SEIR model with reactive contact tracing and social distancing/lockdown (self-quarantine) measures. The model variables include: susceptible ($S$), exposed ($E$) and infectious ($I$) individuals; social-distanced (or self-quarantined) susceptible ($S_q$), exposed ($E_q$) and infectious ($I_q$) individuals; contact-traced susceptible ($S_c$), exposed ($E_c$), and infectious ($I_c$) individuals; and the decoupled compartments of (safely) isolated reported cases ($R$) with a subset of currently quarantined contact-traced cases ($R_c$).

***Full Model:***

$$\begin{aligned}
S' &= -S\left(1+\psi\right)\lambda + (1-\theta_c)\alpha_c S_c + \alpha_q S_q \\
(S_c)' &= S\frac{(1-p)}{p}\phi\lambda + \frac{(1-p)}{p}\nu_q\phi_q S_q\lambda - \alpha_c S_c - \nu_c S_c\lambda \\
(S_q)' &= \sigma S\lambda - \alpha_q S_q + \theta_c\alpha_c S_c - \nu_q\left(1+\frac{(1-p)}{p}\phi_q\right)S_q\lambda \\
E' &= \left(1-\phi-\xi\right)S\lambda + \left(1-\phi_c-\xi_c\right)\nu_c S_c\lambda + \left(1-\phi_q-\xi_q\right)\nu_q S_q\lambda - \frac{1}{\tau}E \\
(E_c)' &= \phi S\lambda + \phi_c\nu_c S_c\lambda + \phi_q\nu_q S_q\lambda - \frac{1}{\tau}E_c \\
(E_q)' &= \xi S\lambda + \xi_c\nu_c S_c\lambda + \xi_q\nu_q S_q\lambda - \frac{1}{\tau}E_q \\
I' &= \frac{1}{\tau}E - \frac{1}{T}I \\
(I_c)' &= \frac{1}{\tau}E_c - \frac{1}{T_c}I_c \\
(I_q)' &= \frac{1}{\tau}E_q - \frac{1}{T_q}I_q \\
R' &= \frac{1}{T}I + \frac{1}{T_q}I_q + \frac{1}{T_c}I_c \\
(R_c)' &= \frac{1}{T_c}I_c - \alpha_c R_c,
\end{aligned} \qquad [1]$$

where $\lambda = \left(\beta I + \beta_q I_q + \beta_c I_c\right)/N$ is "force of infection", $\psi = \frac{1-p}{p}\phi + \sigma$ is total (contact-traced and self-) quarantine factor and $C_q = S_c + E_c + I_c + R_c$ is the number of quarantined contacts at time $t$. The description of all variables and parameters for the model Eq. (1) are given in Table S1. For model fitting, we take $\phi = \phi_q, \phi_c = 1, \xi = \xi_c = 0, \xi_q = 1-\phi_q$, which specifies that the proportion of non-quarantined and quarantined susceptible individuals contact-traced upon infection is uniform, while all contact-traced susceptible individuals continue to be traced and quarantined susceptibles whom are not traced remain quarantined upon infection (see Fig. 1 in main text for the schematic diagram). For deriving final size results in next section, we specify different conditions for these parameters, which serve as approximations of the final size in the general model, along with exact formulae for important special cases which do not require some of these parameters, e.g. "perfectly shielding" quarantine. Then this fully simplified model (including decouple compartments of contact-traced which are fit to data): Then this simplified model (equation (1) in main text), including the decoupled compartments of contact-traced that are fit to data and self-quarantined susceptibles, becomes:

$$\begin{aligned}
&S' = -\left(1+\frac{(1-p)}{p}\phi+\sigma\right)\beta SI/N, \quad E' = \left(1-\phi\right)\beta SI/N - \frac{1}{\tau}E, \\
&I' = \frac{1}{\tau}E - \frac{1}{T}I, \quad R' = \frac{1}{T}I + \frac{1}{T_c}I_c, \\
&(S_c)' = \frac{(1-p)}{p}\phi\beta SI/N - \alpha_c S_c, \quad (E_c)' = \phi\beta SI/N - \frac{1}{\tau}E_c, \\
&(I_c)' = \frac{1}{\tau}E_c - \frac{1}{T_c}I_c, \quad (R_c)' = \frac{1}{T_c}I_c - \alpha_c R_c, \quad (S_q)' = \sigma\beta SI/N
\end{aligned} \qquad [2]$$

**B. Reproduction number and final size.** The *time-dependent* effective reproduction number $\mathcal{R}_t$ can be defined utilizing the next-generation approach (1). First define the feasible region for the system Eq. (1) as

$$\Gamma = \left\{\mathbf{x} = \left(S, S_c, S_q, E, E_c, E_q, I, I_c, I_q\right)^T \in \mathbb{R}^9_+ \mid N := \sum_{i=1}^{9}\mathbf{x}_i \le N_0\right\},$$

where $\mathbb{R}^9_+$ denotes the non-negative orthant of $\mathbb{R}^9$ and $N_0$ is the initial total population size (of all compartments). We note that the system Eq. (1) is quasi-positive, and thus its solutions remain non-negative when their initial values are nonnegative.

Summing the right-hand sides of Eq. (1), we find that $N'(t) = 0$. Thus the solutions the system Eq. (1) remain in $\Gamma$ when their initial values are in $\Gamma$. Notice that any susceptible population distributed among the defined classes, $S, S_c, S_q$, $\mathcal{E}_0 := (S, S_c, S_q, 0, 0, 0, 0, 0)^T$ is a disease-free equilibrium of system Eq. (1).

We define a next-generation matrix by considering the linearized system at the disease-free equilibrium, $\mathcal{E}_0$. Write the linearized "infection" sub-system as $\mathbf{y}' = (F - V)\mathbf{y}$, where $F$ contains entries corresponding to new infections, and $-V$ contains all other transition terms in the Jacobian matrix evaluated at $\mathcal{E}_0$. Define the current susceptibility and infection transition probabilities of the population by $\mathcal{S} = (1-\phi-\xi)\, S + (1-\phi_c-\xi_c)\,\nu_c S_c + (1-\phi_q-\xi_q)\,\nu_q S_q$, $\mathcal{S}^c = \phi S + \phi_c \nu_c S_c + \phi_q \nu_q S_q$ and $\mathcal{S}^q = \xi S + \xi_c \nu_c S_c + \xi_q \nu_q S_q$. Thus, we consider the following matrices:

$$F = \begin{pmatrix} 0 & 0 & 0 & \mathcal{S}\beta/N & \mathcal{S}\beta_c/N & \mathcal{S}\beta_q/N \\ 0 & 0 & 0 & \mathcal{S}^c\beta/N & \mathcal{S}^c\beta_c/N & \mathcal{S}^c\beta_q/N \\ 0 & 0 & 0 & \mathcal{S}^q\beta/N & \mathcal{S}^q\beta_c/N & \mathcal{S}^q\beta_q/N \\ 0 & 0 & 0 & 0 & 0 & 0 \\ 0 & 0 & 0 & 0 & 0 & 0 \\ 0 & 0 & 0 & 0 & 0 & 0 \end{pmatrix}, \quad V = \begin{pmatrix} \frac{1}{\tau} & 0 & 0 & 0 & 0 & 0 \\ 0 & \frac{1}{\tau} & 0 & 0 & 0 & 0 \\ 0 & 0 & \frac{1}{\tau} & 0 & 0 & 0 \\ -\frac{1}{\tau} & 0 & 0 & 1/T & 0 & 0 \\ 0 & -\frac{1}{\tau} & 0 & 0 & 1/T_c & 0 \\ 0 & 0 & -\frac{1}{\tau} & 0 & 0 & 1/T_q \end{pmatrix}.$$

The next-generation matrix describing expected number of new infections (by the different types of infectious cases) is then defined as $FV^{-1}$. The effective reproduction number, $\mathcal{R}_e$, is the spectral radius $\varrho(FV^{-1})$:

$$\mathcal{R}_e = \varrho(FV^{-1}) = \beta T \mathcal{S} + \beta_c T_c \mathcal{S}^c + \beta_q T_q \mathcal{S}^q \tag{3}$$

Next we derive the following theorem about asymptotic behavior and final size of an outbreak in our model.

**Theorem 1.** *Consider model* Eq. (1) *with non-negative initial conditions satisfying* $\beta(E(t_0)+I(t_0)) + \beta_q(E_q(t_0)+I_q(t_0)) + \beta_c((E_c(t_0)+I_c(t_0)) > 0$. *Then all variables remain non-negative and* $\lim_{t\to\infty}(E(t_0)+I(t_0)+E_q(t_0)+I_q(t_0)+(E_c(t_0)+I_c(t_0)) = 0$. *Next suppose that* $\phi = \phi_c = \phi_q$, $\xi = \xi_c = \xi_q$, $\alpha_q = 0$, $(1-\theta_c)\alpha_c = 0$. *In addition let* $\nu_c = \nu_q$, *and denote* $\nu = \nu_c = \nu_q$ *and* $S_m = S_c + S_q$. *Define the final (cumulative) epidemic size* $\mathcal{C}_\infty$, *and the final proportion of susceptible (not monitored) individuals* $U_\infty := \frac{S(\infty)}{S(t_0)}$. *Then the following final size formula holds:*

$$\begin{aligned} \ln(U_\infty) &= (1+\psi)\mathcal{R}_e \left( U_\infty - 1 - \frac{S_m(t_0)}{S(t_0)} \left( U_\infty^{\nu/(1+\psi)} + 1 \right) + \frac{\psi}{1+\psi-\nu} \left( (U_\infty)^{\nu/(1+\psi)} - U_\infty \right) \right) \\ &\quad - (1+\psi)\left[ \beta T (E(t_0)+I(t_0))/N + \beta_q T_q (E_q(t_0)+I_q(t_0))/N + \beta_c T_c (E_c(t_0)+I_c(t_0))/N \right], \\ \mathcal{C}_\infty &= S(t_0) \left( \frac{N}{S(t_0)} - \frac{\psi}{1+\psi-\nu} \left( \frac{1-\nu}{\psi} U_\infty + U_\infty^{\nu/(1+\psi)} \right) \right) - S_m(t_0) U_\infty^{\nu/(1+\psi)}. \end{aligned} \tag{4}$$

*Proof.* Non-negativity and boundedness of solutions has already been demonstrated. Now, inspired by final size derivation in (2), we write the model as follows:

$$\begin{aligned} x' &= \pi D y \beta b x - V x \\ y' &= -D y \beta b x + g(y) + A y, \quad \text{where} \end{aligned} \tag{5}$$

$$D = \begin{pmatrix} 1 & 0 & 0 \\ 0 & \nu_c & 0 \\ 0 & 0 & \nu_q \end{pmatrix}, \; b^T = \begin{pmatrix} 0 \\ 0 \\ 0 \\ 1/N \\ \frac{\beta_c}{N\beta} \\ \frac{\beta_q}{N\beta} \end{pmatrix}, \; \pi = \begin{pmatrix} 1-\phi-\xi & 1-\phi_c-\xi_c & 1-\phi_q-\xi_q \\ \phi & \phi_c & \phi_q \\ \xi & \xi_c & \xi_q \\ 0 & 0 & 0 \\ 0 & 0 & 0 \\ 0 & 0 & 0 \end{pmatrix},$$

$$g(y) = S \begin{pmatrix} -\psi \\ \frac{1-p}{p}\phi \\ \sigma \end{pmatrix} + \nu_q S_q \begin{pmatrix} 0 \\ \frac{1-p}{p}\phi_q \\ -\frac{1-p}{p}\phi_q \end{pmatrix}, \quad A = \begin{pmatrix} 0 & (1-\theta_c)\alpha_c & \alpha_q \\ 0 & -\alpha_c & 0 \\ 0 & 0 & \alpha_q \end{pmatrix}.$$

Then

$$\begin{aligned} &(x+\pi y)' = -Vx + \pi g(y(t)) x(t) S(t) + \pi A y \\ &x(t_0) - x(\infty) + \pi(y(t_0) - y(\infty)) = V \int_{t_0}^{\infty} x(t)dt + \int_{t_0}^{\infty} \pi g(y(t)) x(t) S(t) dt + \int_{t_0}^{\infty} \pi A y(t) dt \\ &\Rightarrow x(t_0) - x(\infty) + \pi(y(t_0) - y(\infty)) + \int_{t_0}^{\infty} \pi A y(t) dt = V \int_{t_0}^{\infty} x(t) dt - \int_{t_0}^{\infty} \pi g(y(t)) x(t) S(t) dt \qquad [6] \\ &\Rightarrow \int_{t_0}^{\infty} x(t) dt = V^{-1}\left[ \pi(y(t_0) - y(\infty)) + x(t_0) \right] + \int_{t_0}^{\infty} \pi g(y(t)) x(t) S(t) dt + \int_{t_0}^{\infty} \pi A y(t) dt. \end{aligned}$$

Note that the integrals on the right-hand side of Eq. (6) can be bounded in norm as $C\int_{t_0}^{\infty}\|x(t)\|\,dt$ for an appropriate positive constant $C$. All other terms in Eq. (6) (including $x(\infty)=\limsup x(t)$) are finite in norm, in particular $\|Ay(t)\|=0$. Therefore the integral $C\int_{t_0}^{\infty}\|x(t)\|\,dt$ is finite, which implies that $\|x(t)\|\to 0$ as $t\to\infty$. This proves the first statement.

Furthermore assuming $\phi=\phi_c=\phi_q$, $\xi=\xi_c=\xi_q$, $\alpha_q=0$, $(1-\theta_c)\alpha_c=0$, then $\pi g(y(t))=\mathbf{0}^T$, and

$$S' = -S(1+\psi)\beta b x$$
$$\Rightarrow \ln\left(\frac{S(t_0)}{S(\infty)}\right) = (1+\psi)\beta b\int_{t_0}^{\infty} x(t)dt$$
$$\Rightarrow \ln\left(\frac{S(t_0)}{S(\infty)}\right) = (1+\psi)\beta b V^{-1}\left(\pi(y(t_0)-y(\infty))+x(t_0)\right) \qquad [7]$$

Now $y(\infty)=(S(\infty),S_c(\infty),S_q(\infty))^T$.

Since $\nu=\nu_c=\nu_q$, we can derive the following relationship between $S$ and $S_m := S_c+S_q$:

$$S_c'+S_q' = -\frac{(1-p)\phi}{p(1+\psi)}S' - \frac{\sigma}{1+\psi}S' - S_c\nu_c\lambda(t) - S_q\nu_q\lambda(t)$$
$$S_m' = -c_1S' + c_2\frac{S'}{S}S_m, \quad \text{where } c_1=\frac{\psi}{1+\psi},\ c_2=\frac{\nu}{1+\psi}$$
$$\Rightarrow \left(S_m(t)S^{-c_2}(t)\right)' = -c_1S'(t)S^{-c_2}(t)$$
$$\Rightarrow S_m(\infty)S^{-c_2}(\infty) - S_m(t_0)S^{-c_2}(t_0) = \frac{c_1}{-c_2+1}\left(S^{-c_2+1}(t_0) - S^{-c_2+1}(\infty)\right) \qquad [8]$$

Define the *final (cumulative) epidemic size* $\mathcal{C}_\infty$, and the final proportion of susceptible (not monitored) individuals $U_\infty := \frac{S(\infty)}{S(t_0)}$. When plugging in Eq. (8) into Eq. (7), we derive the following relationship:

$$\ln(U_\infty) = (1+\psi)\mathcal{R}_e\left(U_\infty - 1 - \frac{S_m(t_0)}{S(t_0)}\left(U_\infty^{\nu/(1+\psi)}+1\right) + \frac{\psi}{1+\psi-\nu}\left((U_\infty)^{\nu/(1+\psi)} - U_\infty\right)\right)$$
$$- (1+\psi)\beta b V^{-1}x(t_0)$$
$$\mathcal{C}_\infty = N - S(\infty) - S_m(\infty),$$
$$\mathcal{C}_\infty = S(t_0)\left(\frac{N}{S(t_0)} - \frac{\psi}{1+\psi-\nu}\left(\frac{1-\nu}{\psi}U_\infty + U_\infty^{\nu/(1+\psi)}\right)\right) - S_m(t_0)U_\infty^{\nu/(1+\psi)},$$

where $N$ is total population size and $\beta bV^{-1}x(t_0) = \beta T(E(t_0)+I(t_0))/N + \beta_qT_q(E_q(t_0)+I_q(t_0))/N + \beta_cT_c(E_c(t_0)+I_c(t_0))/N$. $\square$

If we start from the beginning of an outbreak, letting $t_0=0$, then

$$\ln(U_\infty) = (1+\psi)\mathcal{R}_0\left(U_\infty - 1 + \frac{\psi}{1+\psi-\nu}\left((U_\infty)^{\nu/(1+\psi)} - U_\infty\right)\right)$$
$$\mathcal{C}_\infty = N\left(1 - \frac{\psi}{1+\psi-\nu}\left(\frac{1-\nu}{\psi}U_\infty + U_\infty^{\nu/(1+\psi)}\right)\right).$$

In the case that $\nu=0$, the formula reduces to:

$$\ln(U_\infty) = \mathcal{R}_0(U_\infty-1), \quad \mathcal{C}_\infty = N\frac{1}{1+\psi}(1-U_\infty),$$

where $\mathcal{R}_0 = \beta(1-\phi)T$ and $I(0)\approx 0$ in this case at the outset of the outbreak.

Furthermore in the above special case, along with the restriction that $\nu=0$, a formula measuring peak infected levels can be derived along the lines of the method outlined in (3). For simplicity, we consider the instance of perfect quarantine ($\beta_q=\beta_c=0$) in model Eq. (2). Define $\mathcal{Y}(t) := E(t)+I(t)$. Then it is not hard to see that $\mathcal{Y}'(t)=0$ when $\mathcal{R}_e(t)=1$, where $\mathcal{R}_e(t)=\mathcal{R}_0\frac{S(t)}{N}$. Let $t_p$ the time of peak (non-quaratined) infected, where $\mathcal{Y}(t_p)=\mathcal{Y}_{peak} := \max_{t>0}\mathcal{Y}(t)$. Then we obtain the following: If we obtain the formula if $\mathcal{Y}(0)=E(0)+I(0)\approx 0$:

$$\frac{d\mathcal{Y}}{dS} = \frac{1}{1+\psi}\left(1-\frac{1}{\mathcal{R}_e(t)}\right)$$
$$\Rightarrow \int_0^{t_p} d(\mathcal{Y}(t)) = \int_0^{t_p}\frac{1}{1+\psi}\left(1-\frac{N}{\mathcal{R}_0S(t)}\right)d(S(t))$$
$$\Rightarrow \mathcal{Y}_{peak} = \frac{N}{(1+\psi)\,\mathcal{R}_{0,b}}\left(\ln\frac{1}{\mathcal{R}_0}+\mathcal{R}_0-1\right). \qquad [9]$$

**C. Data Fitting to Model.** We utilize data on total reported cases and quarantined contacts in mainland China published in publicly available daily reports by NHC (National Health Commission of the People's Republic of China) (4). We first fit both our full model Eq. (1) (parameter assumptions in Sect. A and diagram in Fig. 1 in main text) and simplest model Eq. (2) simultaneously to (cumulative) reported case data and (daily number of) quarantined contacts utilizing a weighted least squares algorithm. We utilized a nonlinear weighted least squares algorithm, minimizing the objective function $J(t) = R(t) + wC_q(t)$, where $w = 0.05$ is a chosen positive weight and the variables $R(t)$ and $C_q(t)$ represent cumulative reported cases and number of quarantined contacts, respectively. The fitting optimization algorithm is implemented in MatLab via the lsqcurvefit function, which utilizes the interior-reflective Newton method.

For the model fits presented in main text, we utilize a baseline reproduction number, $\mathcal{R}_{0,b} = 6$, fixed infectious periods and incubation periods. The fixed parameters are detailed in Table S1, and fitted parameters for simplified model ((1) in main text) and full model are presented in Table S1 and Table S3, respectively. Furthermore we conduct an uncertainty analysis detailed in next subsection for the simplified model fit. We note that our model initiates on January 21, 2020, when the data for reported cases begins (with the fitted initial infected $I_0 = 778$ for the simplified model). The data for quarantined contacts begins on Jan. 26. As remarked in the main text, our model actually allows for a very similar epidemic trajectory when initiating the simulation a month earlier with one infected individual ($I_0 = 1$) and all other parameters the same as our fit starting from Jan. 21 (see Fig. S1).

There are possible issues with the case data as detailed by other researchers (5); most notably a change in case counting procedures on Feb. 12 in Hubei province causing an abrupt decrease then sharp increase in reported cases. Although utilizing all cumulative cases reported in China was desirable for model fitting, the data discrepancies, along with potential statistical issues, motivate us to comprehensively test robustness of our results. First, because fitting of the model to cumulative incidence data leads to inconsistent assumptions on independence of errors and possible bias (6), we fit the model to the actual daily incidence (inferred from cumulative case data) and to the quarantined contact data as before. Second, the outbreak was not localized to a single population during the timeframe considered, therefore we test the effects of spatial aggregation on parameter estimates. Due the case counting issues in Hubei around Feb. 12, we excluded the daily incidence numbers for this province (also when fitting incidence for all China) on Feb. 11-13 from our data fitting. We also tested smoothing the data around Feb. 12 and fitting the cumulative cases. Finally, we conducted various other explorations of different modeling assumptions including (i) investigation of susceptible self-quarantine rates dependent on mobility data or proportional to rate of reported cases (instead of force of infection), (ii) allowing more general residence time distributions for quarantine periods, and incubation and infectious periods, (iii) utilizing different baseline reproduction numbers or including unreported cases in the model. All of these additional modeling exercises are summarized below in the following subsections.

***C.1. Spatially Aggregated Daily Count (DC) Fits.*** For spatially aggregated we considered all of China, China less Hubei Province, and Hubei province. For all of these fits the data used were daily case totals, inferred from cumulative case totals, and nationally aggregated quarantine data. To obtain the fit for China less Hubei and Hubei in this circumstance we simultaneously fit their respective case data and the sum of their respective quarantine model compartments with initial conditions chosen where appropriate based upon initial relative reported cases and under the assumption that the probability of transmission given contact, $p = .06$, in line with other studies (7, 8). The results of these fittings are summarized in table S5 and figure S8, along with figure 2 in the main text.

***C.2. Spatially Segregated Provincial DC Fits.*** To test robustness with respect to the effects of spatial aggregation on estimated parameter values, for each province we simultaneously fit model Eq. (2) to daily case incidence and an inferred number of quarantined individuals for that province. Note that we do not include the provinces of Tibet, which had only one confirmed case, and Hong Kong, where the peak daily case total occurred well after the time frame considered. Furthermore because quarantined contact data was only available aggregated for all of China (from NHC (4)), we estimated the number of quarantined individuals for each province, labeled $j = 1, \ldots :, 30$, as follows:

$$\begin{cases} Q_j(0) = \left(\frac{C_j(0)}{C(0)}\right) Q(0) \\ Q_j(t) = \left(\frac{\int_0^t e^{-s/14} C_j(s) ds}{\int_0^t e^{-s/14} C(s) ds}\right) Q(t), \quad t > 0, \end{cases}$$

where $Q_j, C_j$ are the provincial quarantine and daily case totals, and $Q, C$ are the national quarantine and daily case totals. Here the assumption is that quarantined contacts are proportional to reported case load, and as specified in our model, quarantine duration is exponentially distributed with mean 14 *days*. Plots for these fits can be found in figures S5 and S6. Fit parameter values are presented in table S4.

By including spatial (provincial) heterogeneity, first observe that the fit parameter values of Hubei are very close to that of China aggregated. This is expected since a significant amount of total cases in China occurred in Hubei. For the remaining provinces with much smaller outbreaks, we obtain good fits that can be largely mimicked by aggregating and fitting cases to China less Hubei (see tables S4, S5). Observe that $\sigma$ and $\phi$ being significantly higher in these other provinces than in Hubei (or the all China fit). Indeed, comparison of the values in tables S4 and S5 suggests that segregating between China Less Hubei and Hubei alone is sufficient to capture this difference. The higher values for $\sigma$ in China Less Hubei are likely explained by the other provinces having the advantage of responding to the outbreak in Hubei, along with their local cases, so that their lockdowns could be enacted faster (with larger magnitude relative to local force of infection). The larger values of contact tracing coverage $\phi$ in China Less Hubei may be attributed to a smaller caseload enabling this relatively resource-intensive control strategy. Despite the heterogeneity, the conclusion that lockdowns (self-quarantine) had the overwhelming influence on the outbreak compared to contact tracing is robust, as noted in the main text and SI through sensitivity analysis on $R_e$, final and peak outbreak size with respect to $\phi$ and $\sigma$.

***C.3. Weekly $\mathcal{R}_e$.*** In addition to computing $\mathcal{R}_0$ by parameter estimation of differential equation models, we utilize an alternative purely statistical approach incorporating both the case and quarantined contact data to infer $\mathcal{R}_e$ and efficacy of contact tracing developed in a prior study of the 2014-2015 Ebola outbreak (9). The method, based on (10), measures $\mathcal{R}_e$ directly from reported case data and estimates of the serial interval (generation time) distribution. We utilize serial interval distributions from a large study of cases and their contacts in Shenzhen, China (7). The serial interval for (untraced) reported cases is taken to be $Gamma(2.29, 0.36)$ resulting in mean 6.29 $days$. The serial interval for infections caused by contact-traced cases is taken to be $Gamma(1.8, 0.5)$ resulting in mean 3.6 $days$. Let $\mathcal{R}_e^{(j)}$ be the daily reproduction number, $\mathcal{R}_{e,n}^{(j)}$ be the daily reproduction number if there was no contact tracing, $U_j$ be the untraced reported cases on day $j$, $T_j$ be the traced reported cases on day $j$, $\pi_j, \omega_j$ be the c.d.f. of serial interval distributions, and $\kappa = \beta_c/\beta$ be the proportion of transmissions caused by traced cases (relative to untraced). Then

$$\mathbb{E}(\mathcal{R}_e^{(j)}) = \frac{1}{U_j + \kappa T_j} \sum_n (\pi_n U_{j+n} + \kappa \omega_n T_{j+n})$$

$$\mathbb{E}(\mathcal{R}_{e,n}^{(j)}) = \frac{1}{U_j + \kappa T_j} \sum_n (\pi_n U_{j+n} + \omega_n T_{j+n})$$

Note that $\kappa = 0$ for the simplified (perfect tracing) model. Since the amount of infected quarantined contacts is not available in the data, we utilize the predicted relative transmission and incidence of contact-traced individuals from our model fit to assess $\mathcal{R}_e$ with and without contact tracing. The results (Fig. 2 in main text) estimate the proportion of reported cases which are traced contacts and reduction of $\mathcal{R}_e$ due to contact tracing.

**D. Uncertainty Quantification.** We used the following method to generate 95% confidence intervals for the selected quantities appearing in tables S2,S5.

1. Simultaneously fit the simplest model to daily case totals for China less Hubei Province, Hubei Province, and national quarantined data as described in the proceeding section.

2. Based on the two fit daily case total curves, and under the assumption that the reporting error is normally distributed and relative in magnitude to the reported total at each data point:

$$y_i = g(x(t_i), \hat{\xi}) + \epsilon_i \qquad\qquad \epsilon_i \sim n(0, y_i \cdot s^2),$$

   where $g$ is the true number of daily cases and $\hat{\xi}$ the set of true parameter values. We generated 10,000 datasets and refit the model to each of them and the original quarantine data simultaneously. For the results in tables S2,S5 a value of $s = .5$ was used because this value causes the synthetic data-sets to cover the original data except for the outliers around February 12, when there was a change in the method of reporting cases in Hubei province (see figure S2).

3. if fitting aggregate case totals sum the results from step 4 to obtain cumulative case numbers

4. Produce scatter plots and correlation values for each of the fit parameters based on these generated data for the cumulative data fitting (see figures S3 and S4 respectively).

5. Arrange the generated values in increasing order and remove the top and bottom 2.5% in order to obtain the desired approximate 95% Confidence Intervals.

### E. Alternative modeling assumptions.

***E.1. Exploration of different self-quarantine rate forms.*** The models considered so far incorporate mass self-quarantine proportional to *force of infection*, e.g. $\sigma\lambda(t)$ in Eq. (2), as a simple proxy for reactionary lockdowns, individual behavior change, etc., occurring population-wide during the outbreak. In reality, this proportionality relationship has several limitations. In particular, there is a delay between new incidence and case reporting which may delay the action of self-quarantine with respect to force of infection. Additionally, while public health proclamations are generally reactionary, other factors may come into play. In order to test the robustness of our simplification we explore different (susceptible) self-quarantine rates here. First, if we simply take self-quarantine rate dependent on the (instantaneous) rate of change in (cumulative) reported case prevalence, $R'(t)/N$, then we obtain the modified factor $\tilde{\sigma}$:

$$\tilde{\sigma} \frac{R'(t)}{N} S(t) = \tilde{\sigma} \left(\frac{1}{T}\right) I(t)S(t) = \sigma\lambda(t) = \sigma \frac{\beta}{N} I(t)S(t) \quad \Rightarrow \quad \tilde{\sigma} = \sigma\mathcal{R}_{0,b}. \qquad [10]$$

Thus, the form of self-quarantine rate is preserved (proportional to $I(t)S(t)$), but the proportionality constant is modified in such a way that the factor must be $\mathcal{R}_{0,b}\times$ larger when relative to rate of reporting to reach compared to the equivalent quarantine levels when proportional to force of infection.

Next, we consider the following delay equation with self-quarantine dependent on daily reported cases, which modifies the susceptible compartment in Eq. (2) as

$$S'(t) = -\frac{\beta}{N}\left(1 + \frac{(1-p)}{p}\phi\right) I(t)S(t) - \tilde{\sigma}\,\frac{(R(t) - R(t-1))}{N}S(t). \qquad [11]$$

Here, the factor proportional to (prevalence of) new reported cases within the past 24 hours, $\tilde{\sigma}(R(t) - R(t-1))/N$, replaces the force of infection relationship ($\sigma\lambda(t)$). In Figure S9, we show simulations of the delay system plotted with the original model output (corresponding to all China daily incidence fit) for different values of $\tilde{\sigma}$ (relative to $\sigma$) in Eq. (11). Notice that the incidence trajectory of original model fit is bounded between outbreak simulations of the delay model corresponding to $\tilde{\sigma} = \mathcal{R}_{0,b}\sigma$ (lower bound) and $\tilde{\sigma} = (\mathcal{R}_{0,b}/2)\sigma$. This is consistent with the predicted magnification of $\sigma$ derived above in Eq. (10), and possibly not exactly matching due to the fact that $(R(t) - R(t-1)) > R'(t)$ at the beginning of the outbreak.

Finally, we utilize Baidu mobility data for within-cities, namely City Movement Intensity (WCMI) (11), as a proxy for the rate of self-quarantine in China during the timeframe. We consider the following modification to the susceptible compartment in Eq. (2)

$$S'(t) = -\frac{\beta}{N}\left(1 + \frac{(1-p)}{p}\phi\right) I(t)S(t) - \tilde{\sigma}(t)S(t). \qquad [12]$$

We consider the form

$$\tilde{\sigma}(t) = \begin{cases} \sigma_m & 0 < t \le t_q \\ 0 & t_q < t, \end{cases}$$

where $\sigma_m$ is the rate and $t_q$ is the duration in exponential decay of mobility fit to the WCMI data separately for Hubei and China less Hubei via non-linear least squares method applied to function $WCMI(t) = (WCMI(0) - b)e^{-\sigma_m t} + b$ (with $b$ as a lower bound for movement intensity). Utilizing this fitted self-quarantine rate $\sigma_m$ in Eq. (12), we fit the remaining model parameters to the model, this time fitting $p$, instead of fixing $p = .06$, with the susceptible compartment as described above. The results are consistent with our original model where $\sigma$ is fitted proportionality constant relative to force of infection (see table S5 and figures S13-15).

***E.2. Residence times of quarantine, exposed and infectious periods.*** We considered two possible alternatives for the distribution of quarantine residence times besides the base assumption of an exponential distribution with mean time of 14 days: a gamma distribution with shape parameter $\alpha$ and scale parameter $\beta$ constrained by $\beta = 14\alpha^{-1}$ so that a mean residency time of 14 days was retained; and a Weibull distribution with shape parameter $\lambda$ and scale parameter $\kappa$ similarly constrained by $\lambda = 14\left[\Gamma(1+\kappa^{-1})\right]^{-1}$, with the unconstrained parameters being determined by nonlinear least squares fit to the data together with $I_0$, $\beta$ (and so $\mathcal{R}_{0,b}$), $p$, $\sigma$, and $\phi$. As can be seen in figure S13 both of these resulted in distributions with similar shape, and resulted in a slight reduction in residual, with the fit Weibull distribution having the lowest residual value. These fit distributions indicate that more individuals spend close to no time in quarantine, presumably as testing results are returned (12), and fewer individuals spend significantly more than 14 days in quarantine as would be indicated by the baseline assumption that quarantine residency time is exponentially distributed. However the fit parameter values themselves are comparable (see table S5).

We additionally considered the possibility of the infectious and exposed residency times following an Erlang distribution via the linear chain trick using the following system of ODEs:

$$\begin{aligned}
S' &= -\left(1+\psi\right)\beta SI/N, \quad E_1' = \left(1-\phi\right)\beta SI/N - \frac{n_e}{\tau}E_1, \\
E_j' &= \frac{n_e}{\tau}\left(E_{j-1} - E_j\right) \quad 2 \le j \le n_e \\
I_1' &= \frac{n_e}{\tau}E_{n_e} - \frac{n_i}{T}I_1, \quad I_k' = \frac{n_i}{T}\left(I_{k-1} - I_k\right) \qquad 2 \le k \le n_i \\
R' &= \frac{n_i}{T}I_{n_i} + \frac{n_i}{T_e}(I_c)_{n_i}, \quad (S_c)' = \frac{(1-p)}{p}\phi\beta SI/N - \alpha_c S_c, \\
(E_c)_1' &= \phi\beta SI/N - \frac{n_i}{\tau}(E_c)_1, \quad (E_c)_j' = \frac{ni}{\tau}((E_c)_{j-1} - (E_c)_j) \quad 2 \le j \le n_e \\
(I_c)_1' &= \frac{n_e}{\tau}(E_c)_{n_e} - \frac{n_i}{T_e}I_c, \quad (I_c)_k' = \frac{n_i}{T_e}((I_c)_{k-1} - (I_c)_k) \quad 2 \le k \le n_i \\
(R_c)' &= \frac{n_i}{T_e}I_c - \alpha_c R_c, \quad I = \sum_{s=1}^{n_i} I_s
\end{aligned} \qquad [13]$$

In order to obtain an upper bound for the number of stages in Eq. Eq. (13) we took the variance and standard deviation for the distributions of $T$ and $T_e$ given in (7) (where they are assumed to be log normal), fixed the mean to be the values given in table S1, and set the variances equal, solved for $n_i$ and rounded up. This process resulted in an upper bound of at most two stages.

With this in mind we considered three cases, $n_i = 2$ and $n_e = 1$, $n_i = 1$ and $n_e = 2$, and finally $n_i = 2$ and $n_e = 2$. The fit parameter values corresponding to these fittings are given in table S5, and associated fitting plots are figures S14-S16. We additionally plotted the log normal distributions given in (7) together with exponential distributions and Erlang distributions with the same mean (figures S17,S18). In figure S19 the considered Erlang and Exponential distributions for $\tau$ are shown. The primary difference in these distributions is that under the exponential assumption a greater proportion of individuals become rapidly infectious and have a shorter infectious period respectively. The primary effects of the Erlang assumption for infectious dwell time are to increase $I_0$, as well as overall case totals, and decrease $\sigma$, as indicated by the fit values in table S5 and the plots in figure S14. In turn an Erlang assumption for time until infectiousness lowers the fit value for $I_0$ and increases $\sigma$ and $\phi$. Simultaneously assuming both dwell times follow an Erlang distribution results in more moderate increases in $I_0$ and $\phi$, as well as a slight decrease in $\sigma$ (See table S5 and figures S15,S16). The assumption that infectious period ($T$) is exponential results in better fits than the case of Erlang distribution, whereas either distribution for time until infectiousness ($\tau$) provides good fits to the data.

Furthermore, we tested how varying the means for the quarantine, exposed and infectious periods affects the fitting results (see Fig. S20 for example output). For the quarantine duration, we vary the (exponential distribution) parameter from 1/20 to 1/4, with 1/14 as our baseline assumption (mean duration of 14 days). The parameter fits and results do not change significantly with the varied mean quarantine duration. For exposed ($\tau$) and infectious periods ($T$ and $T_c$) under the assumption of exponential durations, we vary $\tau$ from 2 to 4, along with varying $T$ and $T_c$ in the reported 95% confidence intervals of (4.13,5.1) and (2.08,3.31), respectively, given in (7). The parameter fits and results are robust to varying means $\tau, T$ and $T_c$.

***E.3. Model with unreported cases.*** In addition, we consider a version of the model which includes unreported cases. Let $\rho$ be the probability a non-quarantined infected individual becomes a reported case (we assume that all contact-traced cases are reported). Then in the simplified model, the equations become:

$$\begin{aligned}
&S' = -(1+\psi)\,\beta SI/N, \quad E' = (1-\phi\rho)\,\beta SI/N - \frac{1}{\tau}E, \\
&I' = \frac{1}{\tau}E - \frac{1}{T}I, \quad R' = \frac{\rho}{T}I + \frac{1}{T_c}I_c, \\
&(S_c)' = \frac{(1-p)}{p}\phi\rho\beta SI/N - \alpha_c S_c, \quad (E_c)' = \phi\rho\beta SI/N - \frac{1}{\tau}E_c, \\
&(I_c)' = \frac{1}{\tau}E_c - \frac{1}{T_c}I_c, \quad (R_c)' = \frac{1}{T_c}I_c - \alpha_c R_c,
\end{aligned} \qquad [14]$$

***E.4. Time variable Serial Interval.*** The final alternative modeling assumption we consider is the possibility of time variable serial interval, as suggested by (13). We do so by modifying Eq. (2) to add additional loss terms due to the lockdown (self-quarantine rate proportional to force of infection) in the Infected and Exposed compartments:

$$\begin{aligned}
I' &= \frac{1}{\tau}E - \frac{1}{\tau}I - \sigma\beta I^2/N \\
E' &= (1-\phi)\beta SI/N - \frac{1}{\tau}E - \sigma\beta EI/N
\end{aligned} \qquad [15]$$

We assume the same (initial) infectious period value $T = 4.64$. Under this alternative assumption we fit Eq. (15) for China less Hubei Province and Hubei Province (see figure S21). The residual for this fitting was similar to the fitting of the base simplified model. Comparing the fit parameter values to the base assumption (see table S5) this results in decreases in both $\sigma$ and $\phi$. It is expected with the decreasing serial interval of infected cases that less (self)-quarantine will be needed to control the disease, but simulations show that the trajectory of total (self)-quarantined is very close to the original (constant exposed and infectious period) model fit (see figure S21). Furthermore, the main result of minimal impact from contact tracing compared to self-quarantine is preserved, along with the observation that $\sigma$ and $\phi$ are relatively larger for China less Hubei than they are for Hubei.

**F. Additional model fitting tests.** While the fitting procedure presented in detail in main text and here does not explicitly include unreported cases, we also checked several additional versions of the model, along with different assumptions with regard to fixing versus fitting some parameters. We tested inclusion of unreported cases (see above for model) for fitting the proportion of reported cases $\rho$. Furthermore, we varied and fit the infectious period $T$, incubation period $\tau$, and transmission rate $\beta$. In addition, we also attempted to correct for possible issues with the case data as detailed by other researchers (5); most notably a change in case counting procedures on Feb. 12 in Hubei province causing an abrupt increase in reported cases. We utilize this raw data for fitting, rather than separated by provinces, since a major novelty of this work is to incorporate data on the quarantined contacts which was compiled solely for the whole of China. We also tested smoothing the data around Feb. 12 and fitting the model, which resulted in slightly different parameter estimates.

Although certain parameter values changed, the qualitative results on how contact tracing and social distancing/lockdown measures affected outbreak size however were robust when utilizing raw or smoothed data, along with other versions of the model. Parameter values, figures and code of other model/data versions tested are deposited in GitHub (https://github.com/jcmacdonald-codesData?tab=projects). To illustrate the impact of inclusion of reporting probability parameter $\rho$, we include a sensitivity analysis and fitting figure (Fig. S24) for an instance where $\mathcal{R}_{0,b}$ was fit but constrained to be bounded by 4, taking the value $\mathcal{R}_{0,b} = 3.38$ and $\mathcal{R}_0 = \mathcal{R}_{0,b}(1-\phi\rho) = 3.29$ where $\rho = 0.5$ and $\phi$ is estimated to be very low at 0.05.

As an another example, we display parameter estimates and residuals, along with the key epidemiological quantities, namely reproduction number $\mathcal{R}_0$ and final outbreak size, for a range of fixed baseline reproduction numbers (without control) $\mathcal{R}_{0,b}$ with or without smoothing data in cumulative case fitting (Fig. S23), and in daily case incidence fitting (Fig. S22). Observe that outbreak size is not substantially altered by underlying $\mathcal{R}_{0,b}$. In particular estimates of $\phi$ and $p$ are correlated with $\mathcal{R}_{0,b}$ (see also in bootstrapped confidence interval procedure) which diminishes any effect of higher contact tracing proportion in reducing outbreak size or $\mathcal{R}_0$. In addition, smoothed cumulative data fitting results in less cases, but does not change the relative impact of self-quarantine ($\sigma$) versus contact tracing ($\phi$). For values of $3 \leq \mathcal{R}_{0,b} \leq 6$, using the spatially segregated daily incidence data, the outbreak size is more noisy than when fitting cumulative cases, however the main conclusions are robust. Indeed, or Hubei the final outbreak size increases, but for China Less Hubei there is the inverse relationship, yet the impact of varying $\mathcal{R}_{0,b}$ does not change the outcome that contact tracing has minimal impact when compared to self-quarantine (lockdown) measures. Finally, we show that residual decreases as a function $\mathcal{R}_{0,b}$, doing so particularly rapidly while the baseline reproduction number is less than 5.

### 2. Supplementary Tables and Figures

**Table S1. Model variables and parameters.**

| Variable/Parameter | Description | Estimation |
|---|---|---|
| **Variables** | | |
| $S(t), E(t), I(t)$ | (Non-quarantined) susceptible, exposed and infectious individuals | |
| $S_q, E_q(t), I_q(t)$ | Self-quarantined (social-distanced) susceptible, exposed and infectious individuals | |
| $S_c(t), E_c(t), I_c(t)$ | Contact-traced susceptible, exposed and infectious individuals | |
| $R(t)$ | Isolated reported cases | |
| $R_c(t)$ | Currently quarantined contact-traced cases | |
| **Parameters** | | |
| $\beta, \beta_q, \beta_c$ | Transmission rate (for unmonitored infectious) | $1.293 \times 10^{-8}$ [1], [2] |
| $\phi, \phi_q \phi_c$ | Proportion of (non-quarantined,self-quarantined, already initially traced) contacts traced (or remaining traced) | [3], $\phi_q = \phi, \phi_c = 1$ |
| $\sigma$ | self-quarantine (social distancing or lockdown) factor | [3] |
| $p$ | probability of transmission upon contact | [3] |
| $\nu_q, \nu_c$ | Reduction in susceptibility for self-quarantined, contact-traced susceptible individuals | [2], 0 |
| $\alpha_q, \alpha_c$ | Rate of exit from self-, contact-traced quarantine | [2], 14 (days) |
| $\theta_c$ | Fraction of susceptible individuals who return to "social-distanced" (self-quarantined) after completing contact-traced protocol | 1 |
| $\tau$ | Average time to infectiousness (includes pre-symptomatic) | 3 (14) |
| $T, T_q, T_c$ | Average infectious period (time to isolation) of non-quarantined, self-quarantined, contact-traced | 4.64, 2.71, 2.71 (7) |
| $S(0) = N$ | Total initial susceptible population [4] | |

[1] $\beta$ fixed so that baseline reproduction number without control is $\mathcal{R}_{0,b} = 6$ similar to (15), or fitted under various assumptions (see Tables S2, S4 & S5).
[2] Fixed at 0 in simplified model, fit in full model (see Table S3).
[3] Fit in full & simplified model (see Tables S2 & S3).
[4] Population or aggregated populations of provinces in China. (16)

**Table S2. Bootstrapped Confidence Intervals for Selected Quantities using the simplified model and aggregate case totals**

| | 95% CI | Point Estimate | Mean | Std. Dev. | ARE |
|---|---|---|---|---|---|
| $I_0$ | (748.222, 927.433) | 778.05 | 818.59 | 48.9654 | 5.97836 |
| $\beta$ | (1.22298, 1.35522) | 1.2931 | 1.28149 | 3.35816 | 2.13529 |
| $\sigma$ | (1136.76, 1361.61) | 1240.44 | 1243.57 | 56.5373 | 3.56125 |
| $\phi$ | (0.349138, 0.457924) | 0.376917 | 0.385411 | 0.0279016 | 5.24143 |
| $p$ | (0.0754581, 0.0960111) | 0.0816546 | 0.0831421 | 0.0048252 | 4.15688 |
| $\mathcal{R}_0$ | (3.36902, 3.80467) | 3.7385 | 3.65122 | 0.118974 | 2.71322 |
| $\mathcal{R}_{0,b}$ | (5.67462, 6.28822) | 6.0 | 5.94612 | 0.155819 | 2.13529 |

**Table S3. Fit Quantities for full Model using aggregate case totals**

| Quantity | Point Estimate |
|---|---|
| $I_0$ | 761.985 |
| $M_c(0)$ | 0.017 |
| $\phi$ | 0.428009 |
| $\theta$ | 2.33172e-14 |
| $\sigma$ | 1238.05 |
| $\nu$ | 2.33213e-14 |
| $\theta_c$ | 0.16 |
| $p$ | 0.0927884 |
| $\alpha$ | 2.33214e-14 |
| $I_c(0)$ | 1.62392e-5 |
| $I_{cc}(0)$ | 293.306 |
| $\mathcal{R}_0$ | 3.43194 |

**Table S4. Fit Parameter Values for Provinces of China using Daily Case totals and approximate Quarantined Individuals for each Province except Hong Kong, due to its peak daily case total occurring significantly after the end of available quarantine data; and Tibet, which had one confirmed case during the outbreak.**

| Province | $I_0$ | $\mathcal{R}_{0,b}$ | $\sigma$ | $\phi$ |
|---|---|---|---|---|
| Anhui | 24.201 | 6.0 | 64210.0 | 0.41338 |
| Beijing | 22.228 | 4.9219 | 59285.0 | 0.60144 |
| Chongqing | 33.107 | 5.3588 | 61349.0 | 0.62185 |
| Fujian | 22.509 | 5.214 | 147520.0 | 0.67986 |
| Gansu | 6.1867 | 5.232 | 318820.0 | 0.58251 |
| Guangdong | 55.286 | 6.0 | 90103.0 | 0.51555 |
| Guangxi | 18.56 | 4.2585 | 203190.0 | 0.64409 |
| Guizhou | 2.0885 | 6.0 | 231250.0 | 0.31685 |
| Hainan | 4.2958 | 6.0 | 60484.0 | 0.49998 |
| Hebei | 3.0933 | 6.0 | 219520.0 | 0.32616 |
| Heilongjiang | 5.4071 | 6.0 | 65261.0 | 0.26693 |
| Henan | 12.373 | 6.0 | 55652.0 | 0.30274 |
| Hubei | 324.8 | 6.0 | 1178.3 | 0.32015 |
| Hunan | 32.838 | 6.0 | 61102.0 | 0.4532 |
| Inner Mongolia | 1.8439 | 6.0 | 379400.0 | 0.45947 |
| Jiangsu | 15.109 | 6.0 | 123090.0 | 0.4181 |
| Jiangxi | 22.875 | 6.0 | 44021.0 | 0.40488 |
| Jilin | 1.767 | 6.0 | 258030.0 | 0.36291 |
| Liaoning | 3.9735 | 6.0 | 302290.0 | 0.40183 |
| Macau | 1.0651 | 6.0 | 54198.0 | 0.67337 |
| Ningxia | 1.789 | 6.0 | 94699.0 | 0.41355 |
| Qinghai | 1.4552 | 6.0 | 275660.0 | 0.64266 |
| Shaanxi | 8.6792 | 6.0 | 148260.0 | 0.48639 |
| Shandong | 16.491 | 6.0 | 173500.0 | 0.47499 |
| Shanghai | 12.5 | 6.0 | 66041.0 | 0.4777 |
| Shanxi | 4.3513 | 6.0 | 244730.0 | 0.44902 |
| Sichuan | 16.749 | 6.0 | 163170.0 | 0.48628 |
| Tianjin | 3.0767 | 6.0 | 123960.0 | 0.44167 |
| Xinjiang | 1.4438 | 5.572 | 343980.0 | 0.37189 |
| Yunnan | 9.0932 | 6.0 | 252570.0 | 0.54209 |
| Zhejiang | 51.731 | 6.0 | 40251.0 | 0.48586 |

**Table S5. Bootstraped Confidence Intervals for Selected Quantities using Daily Case Totals.** $\sigma_m$ **is the exponential rate of mobility decay.**

| | Parameter | 95% CI | Point Est. | Mean | Std. Dev. | ARE |
|---|---|---|---|---|---|---|
| **China Less Hubei** [1] | $I_0$ | (304.06, 568.13) | 393.08 | 396.12 | 63.401 | 11.407 |
| | $\mathcal{R}_{0,b}$ | (4.3009, 6.0) | 6.0 | 5.7742 | 0.47993 | 3.763 |
| | $\sigma$ | (100100.0, 127610.0) | 114070.0 | 113240.0 | 7011.4 | 4.9488 |
| | $\phi$ | (0.35645, 0.63956) | 0.58842 | 0.53327 | 0.073158 | 11.423 |
| $\sigma_m$ from mobility data | $I_0$ | – | 457.61 | – | – | – |
| | $\mathcal{R}_{0,b}$ | – | 5.8033 | – | – | – |
| | $\phi$ | – | 0.59027 | – | – | – |
| | $p$ | – | 0.050985 | – | – | – |
| | $\sigma_m$ | – | 0.1051 | – | – | – |
| time variable serial interval [1] | $I_0$ | – | 443 | – | – | – |
| | $\mathcal{R}_{0,b}$ | – | 6.0 | – | – | – |
| | $\sigma$ | – | 77068 | – | – | – |
| | $\phi$ | – | 0.37 | – | – | – |
| **Hubei**[1] | $I_0$ | (410.92, 648.33) | 487.2 | 516.21 | 60.431 | 10.547 |
| | $\mathcal{R}_{0,b}$ | (5.7131, 6.0) | 6.0 | 5.9804 | 0.082126 | 0.32743 |
| | $\sigma$ | (1112.9, 1406.2) | 1260.1 | 1249.0 | 74.413 | 4.7559 |
| | $\phi$ | (0.27846, 0.40932) | 0.32135 | 0.33809 | 0.034025 | 9.2804 |
| $\sigma_m$ from mobility data | $I_0$ | – | 487.2 | – | – | – |
| | $\mathcal{R}_{0,b}$ | – | 6.0 | – | – | – |
| | $\phi$ | – | 0.32135 | – | – | – |
| | $p$ | – | .06 | – | – | – |
| | $\sigma_m$ | – | 0.1859 | – | – | – |
| time variable serial interval [1] | $I_0$ | – | 644.44 | – | – | – |
| | $\mathcal{R}_{0,b}$ | – | 6.0 | – | – | – |
| | $\sigma$ | – | 699.03 | – | – | – |
| | $\phi$ | – | 0.21 | – | – | – |
| **China**[1] | $I_0$ | (745.83, 809.05) | 787.55 | 773.52 | 17.143 | 2.2695 |
| | $\mathcal{R}_{0,b}$ | (5.9288, 6.0) | 6.0 | 5.9955 | 0.023302 | 0.075615 |
| | $\sigma$ | (20906.0, 25382.0) | 23495.0 | 23186.0 | 1147.4 | 3.9386 |
| | $\phi$ | (0.32521, 0.4018) | 0.36897 | 0.3637 | 0.019656 | 4.3037 |
| Gamma Quarantine Assumption [1] | $I_0$ | – | 810.733 | – | – | – |
| | $\mathcal{R}_{0,b}$ | – | 6.0 | – | – | – |
| | $\sigma$ | – | 23463.0 | – | – | – |
| | $\phi$ | – | 0.37296 | – | – | – |
| Weibull Quarantine Assumption [1] | $I_0$ | – | 812.079 | – | – | – |
| | $\mathcal{R}_{0,b}$ | – | 6.0 | – | – | – |
| | $\sigma$ | – | 23486.1 | – | – | – |
| | $\phi$ | – | 0.37309 | – | – | – |
| Infectious Erlang Assumption only [1] | $I_0$ | – | 1259.6 | – | – | – |
| | $\mathcal{R}_{0,b}$ | – | 6.0 | – | – | – |
| | $\sigma$ | – | 18118.0 | – | – | – |
| | $\phi$ | – | 0.33983 | – | – | – |
| Exposed Erlang Assumption only [1] | $I_0$ | – | 732.18 | – | – | – |
| | $\mathcal{R}_{0,b}$ | – | 6.0 | – | – | – |
| | $\sigma$ | – | 30887.0 | – | – | – |
| | $\phi$ | – | 0.50246 | – | – | – |
| Simultaneous Erlang Assumption [1] | $I_0$ | – | 1053.1 | – | – | – |
| | $\mathcal{R}_{0,b}$ | – | 6.0 | – | – | – |
| | $\sigma$ | – | 20864.0 | – | – | – |
| | $\phi$ | – | 0.42596 | – | – | – |

[1] $p$ fixed at 0.06

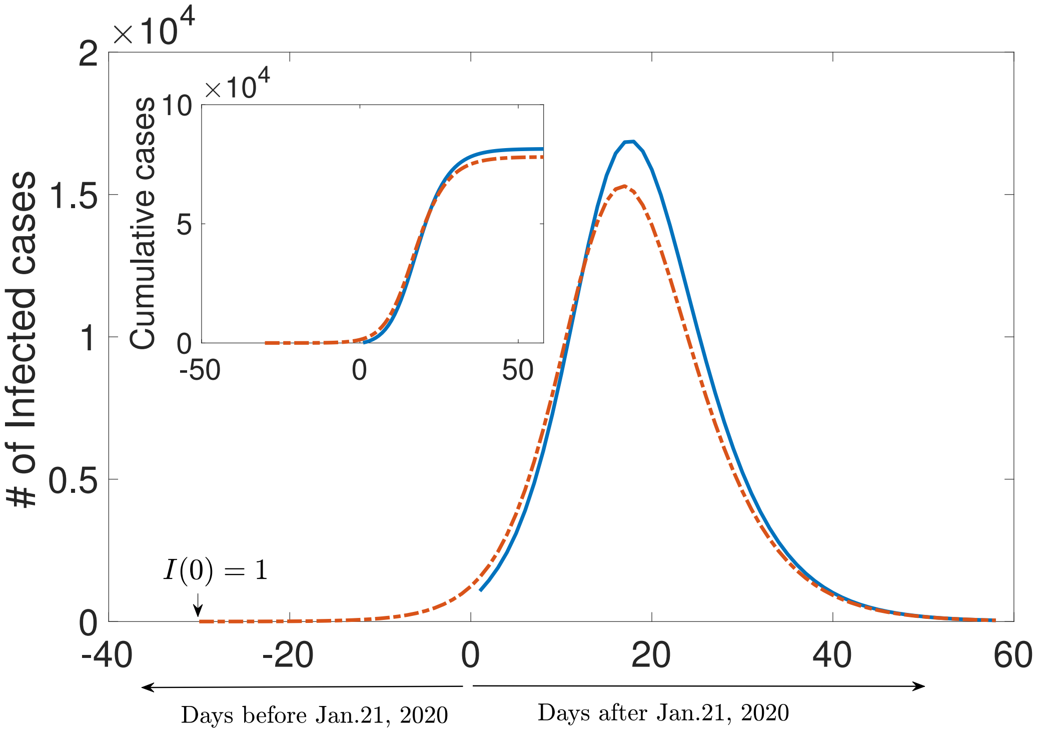


**Fig. S1.** Fitted trajectory starting Jan. 21 (solid blue) alongside simulation with same parameters except initial infected $I_0 = 1$ (dashed red).

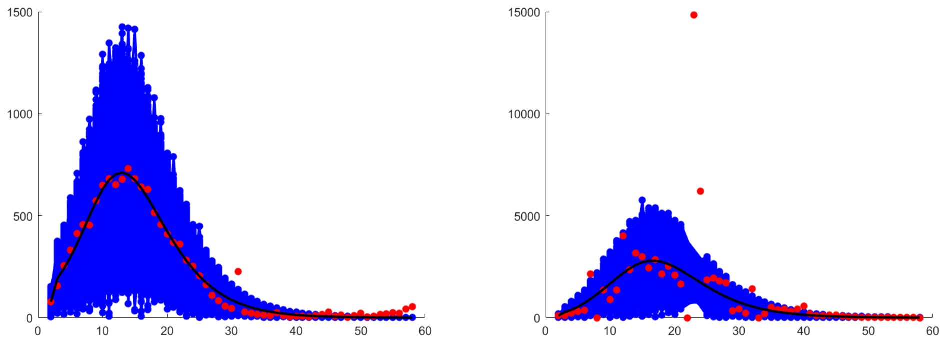

**Fig. S2.** Daily Case totals China Less Hubei (left) and Hubei (right). Synthetic datasets are in blue. Outliers around Day 23 in the Hubei Dataset are excluded from the fit

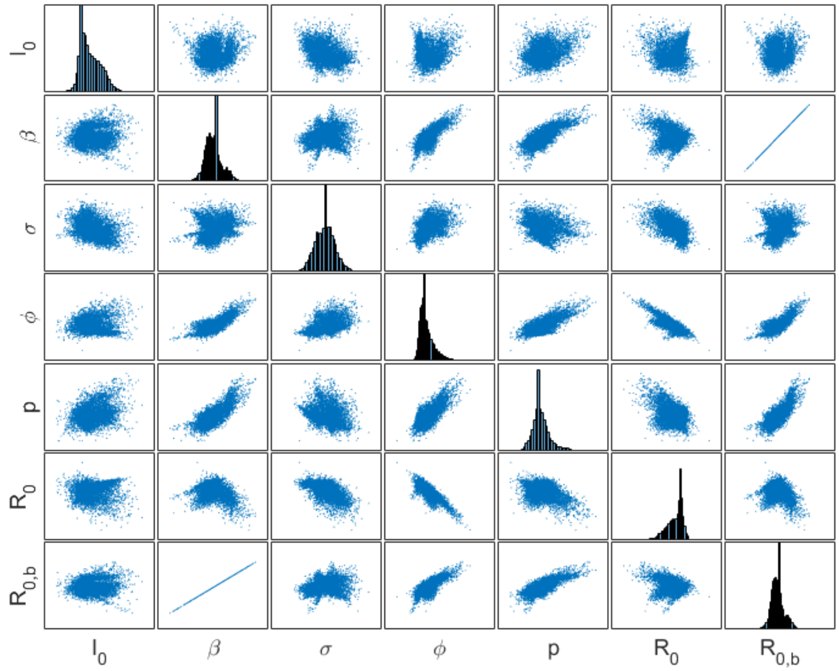


**Fig. S3.** Scatter Plots for selected quantities with histograms on the main diagonal. The relationship between $\mathcal{R}_{0,b}$ and $\beta$ is perfectly linear because $T$ is fixed.

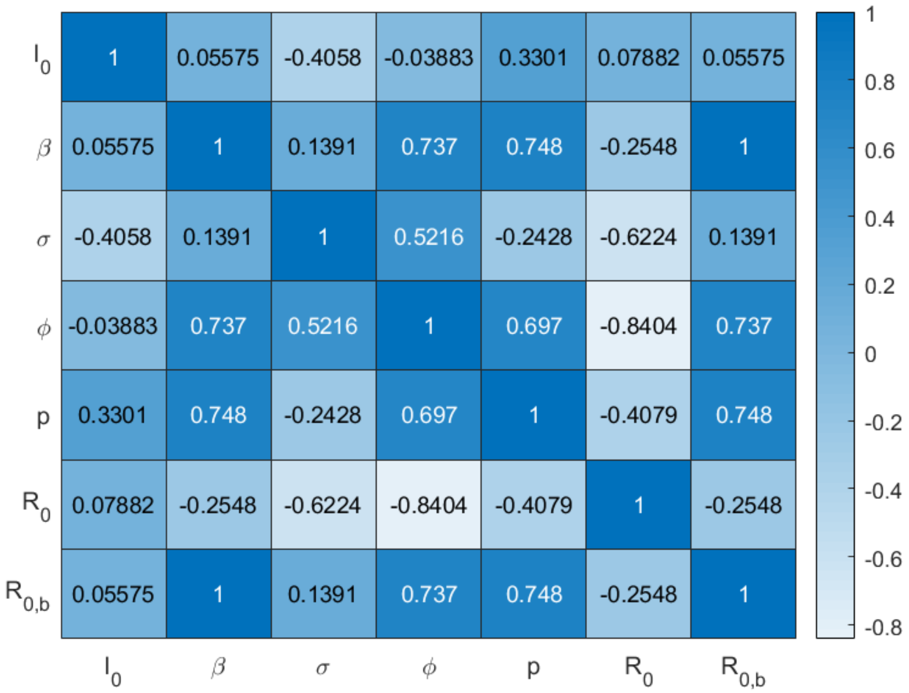


**Fig. S4.** Correlation Values for quantities of interest

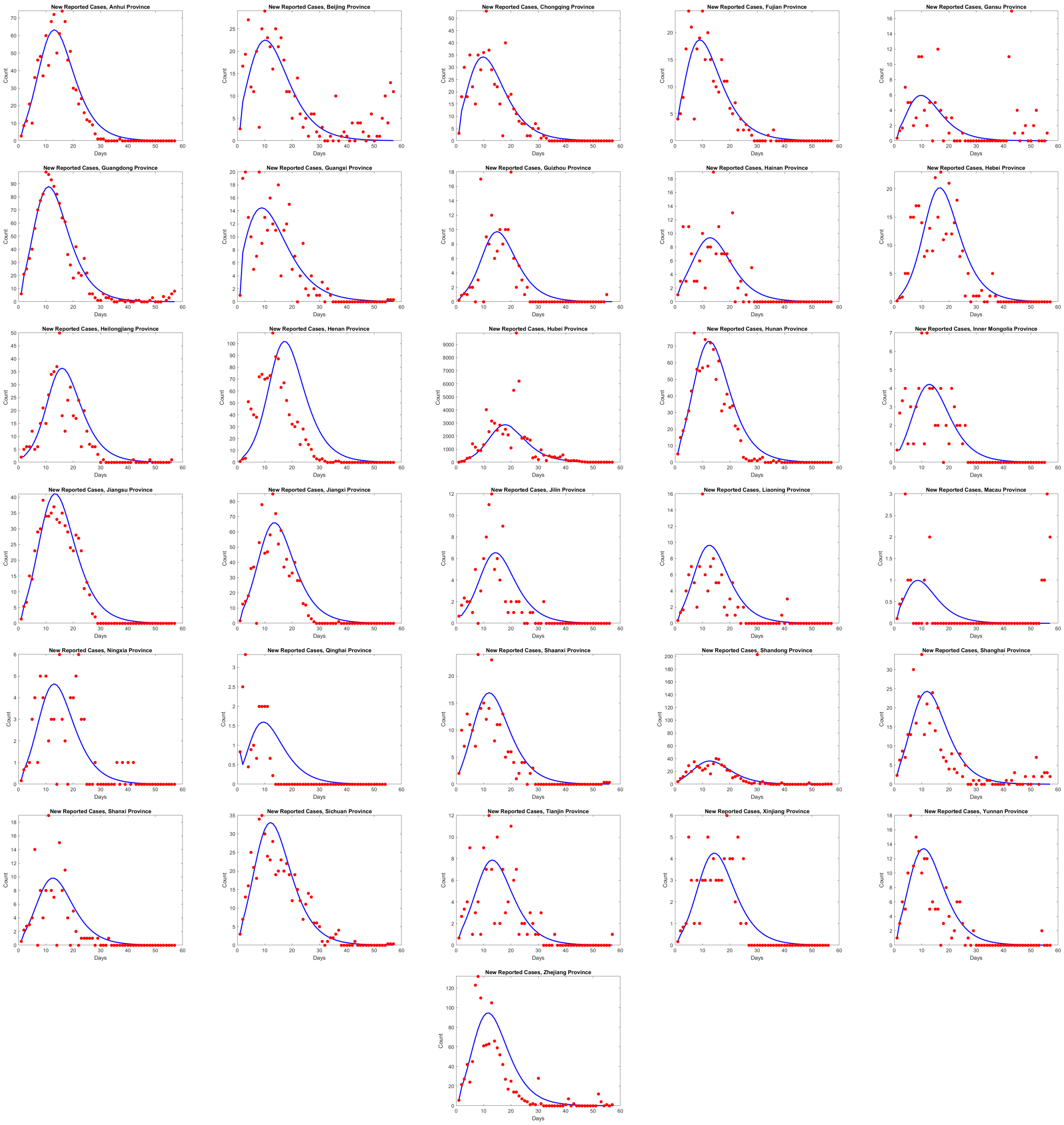


**Fig. S5.** Provincial New Daily Cases fit corresponding to values in table S4

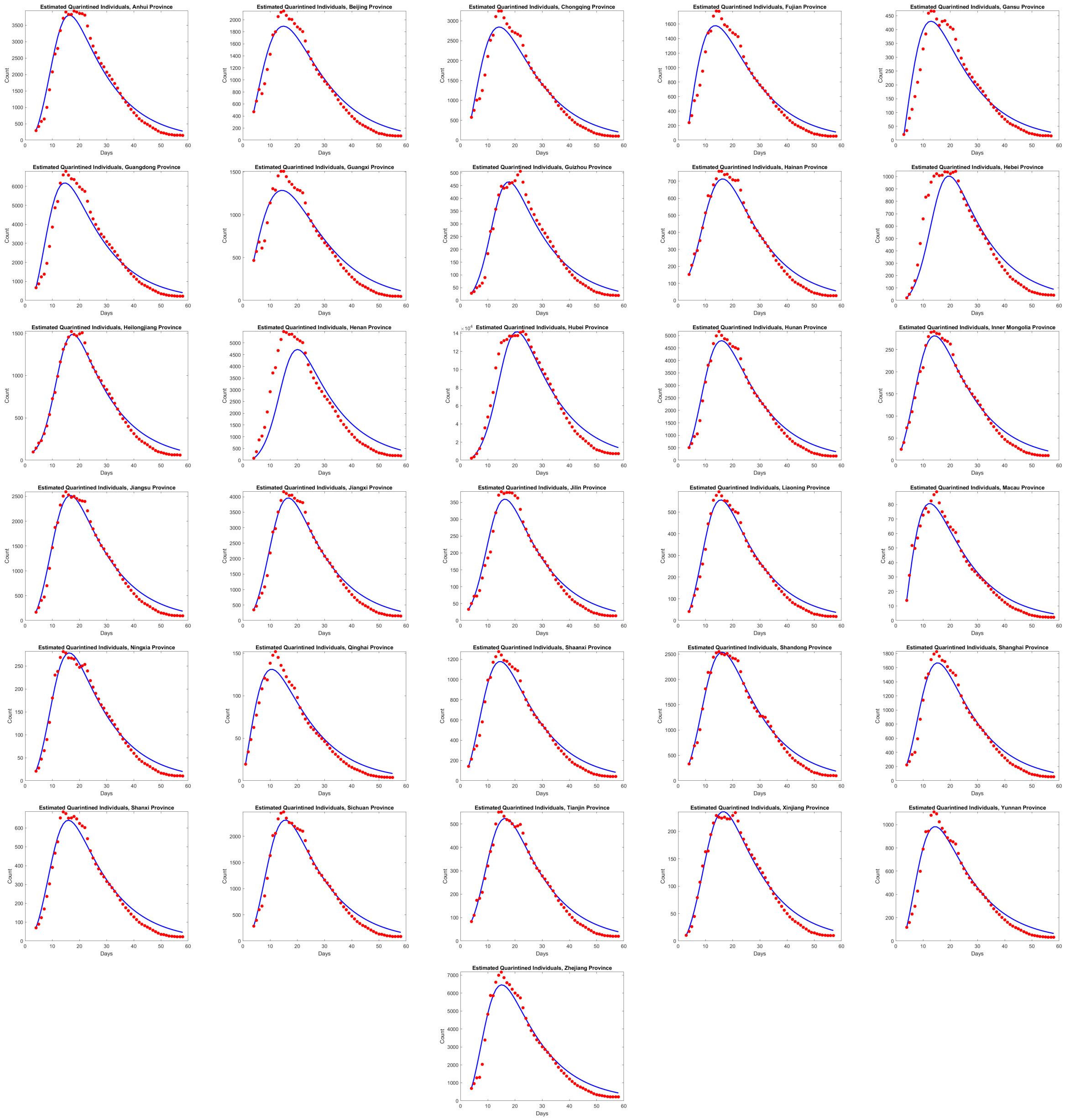


**Fig. S6.** Approximated Provincial Quarantine Data fit corresponding to values in table S4

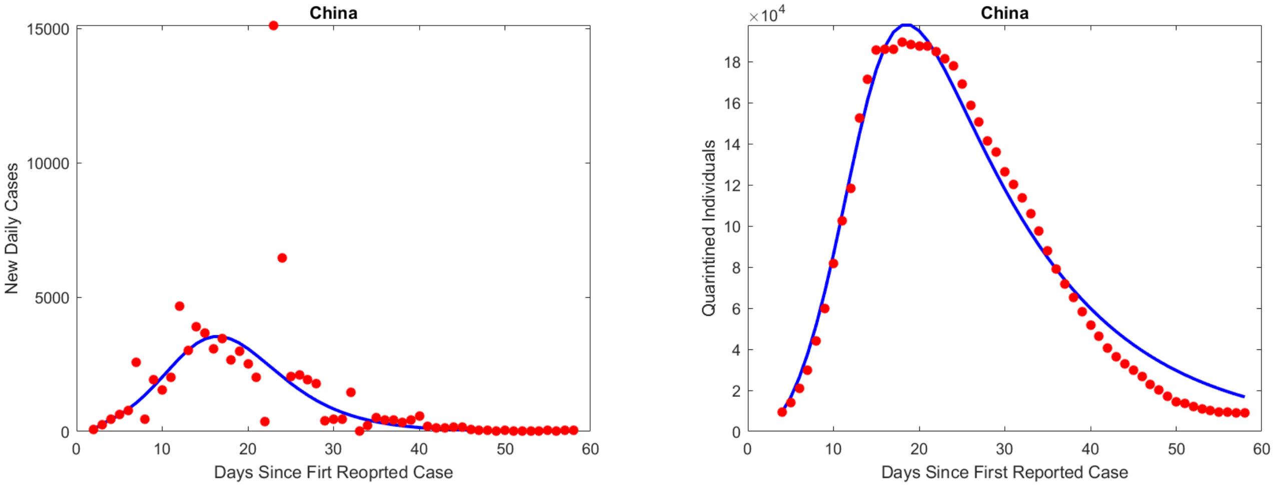


**Fig. S7.** Spatially Aggregated fits corresponding to values in table S5

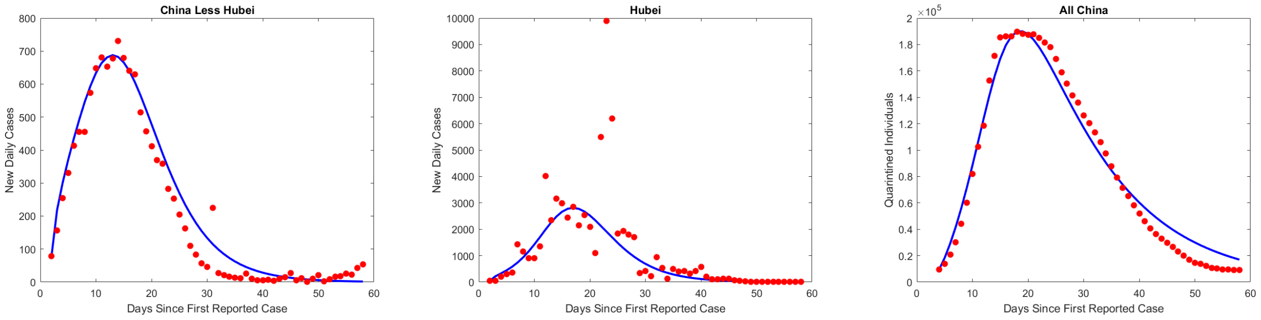


**Fig. S8.** Spatially segregated fits corresponding to values in table S5

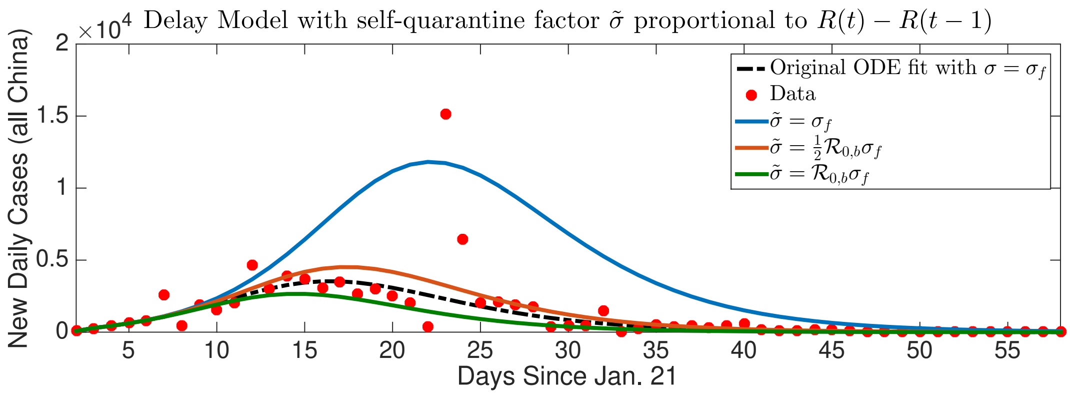


**Fig. S9.** Fitted daily case incidence trajectory in original ODE model (dashed black) alongside simulations with modified self-quarantine proportionality constant $\tilde{\sigma}$ relative to daily reported case variable $(R(t) - R(t-1))$ in DDE model Eq. (11).

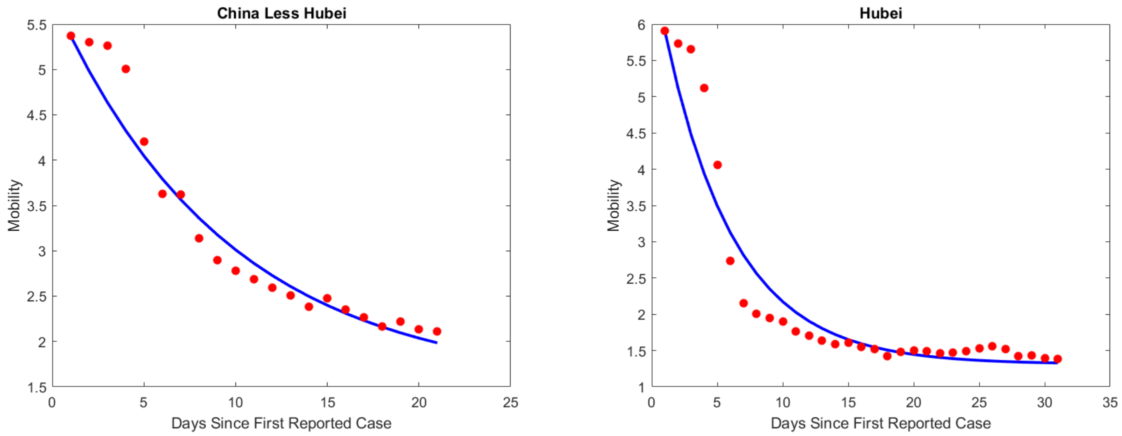


**Fig. S10.** Exponential decay fit using Baidu (WCMI) mobility data to approximate $\sigma_m$, see also table S5.

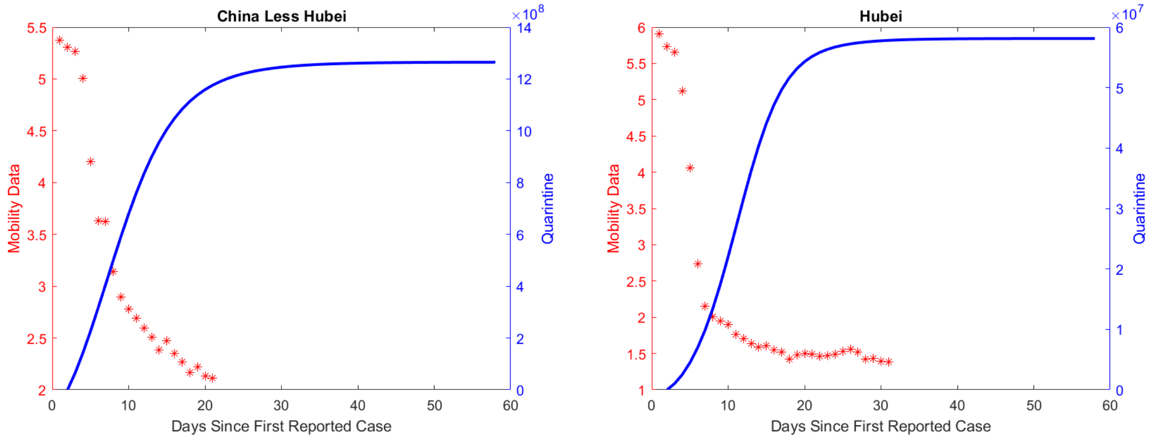


**Fig. S11.** Total self-quarantined trajectory in model fit, alongside Baidu (WCMI) mobility data, see also table S5 and Eq. (12).

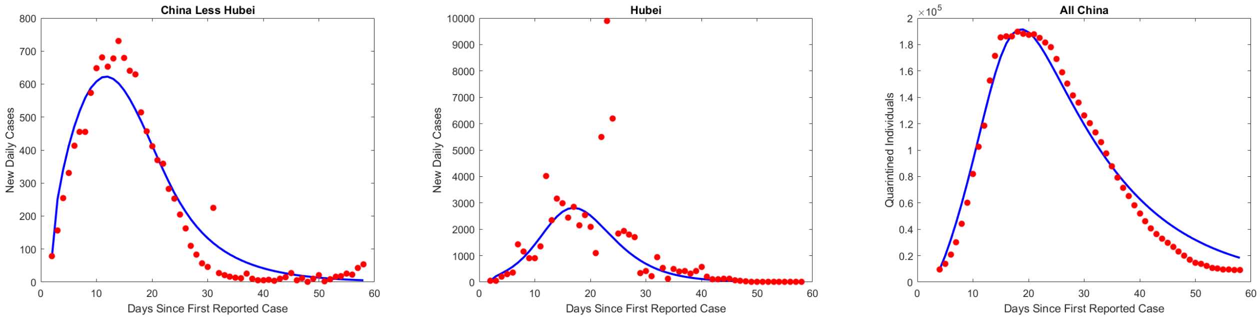


**Fig. S12.** Model fit using mobility data to approximate $\sigma_m$, see also table S5 and Eq. (12).

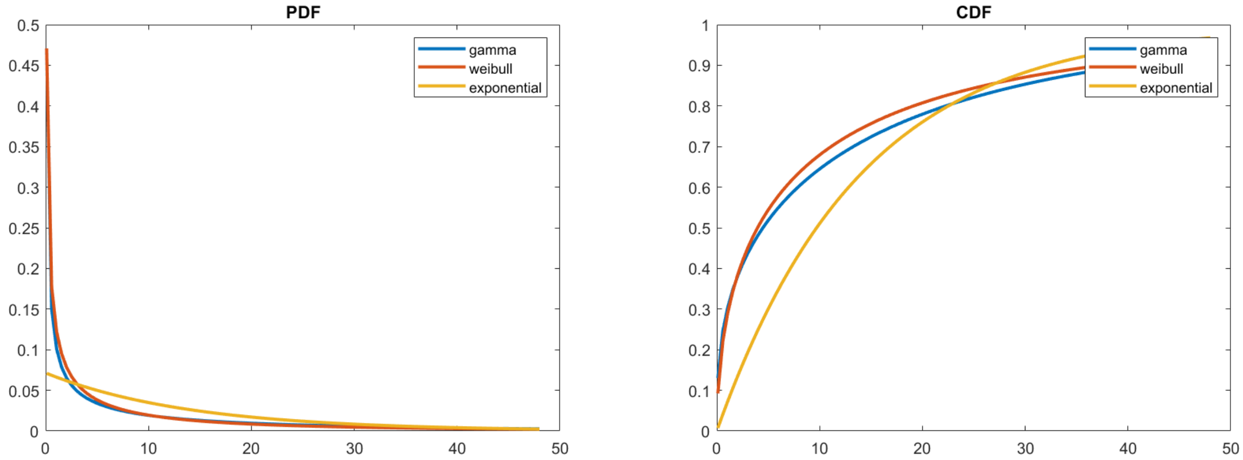


**Fig. S13.** Fit gamma and Weibull distribution curves vs an exponential distribution, each with mean of 14 days, see also table S5

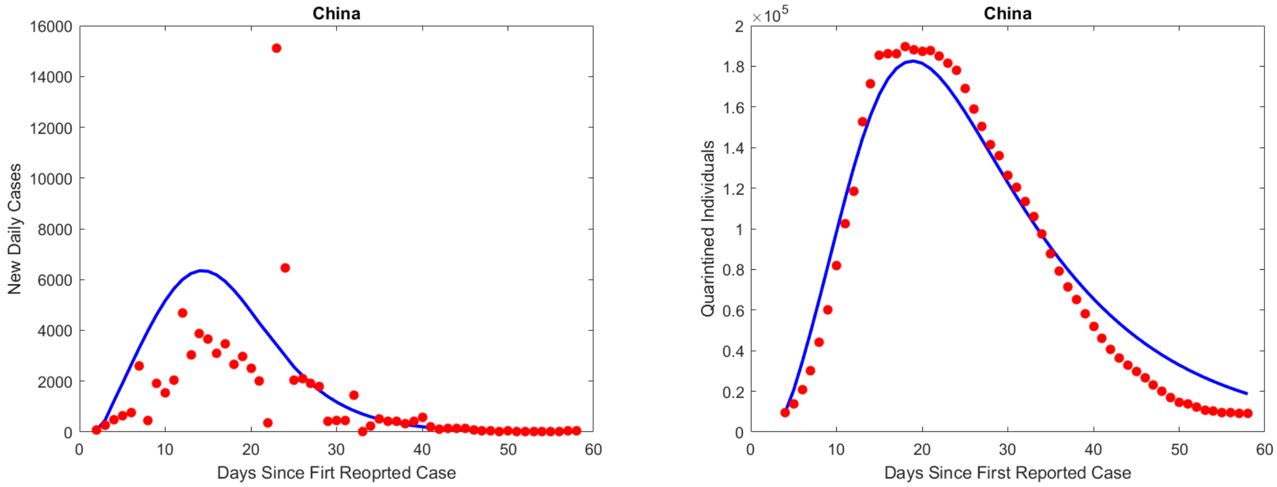


**Fig. S14.** Model fit under the assumption that $T$, $T_e$ follow an Erlang Distribution, see also table S5

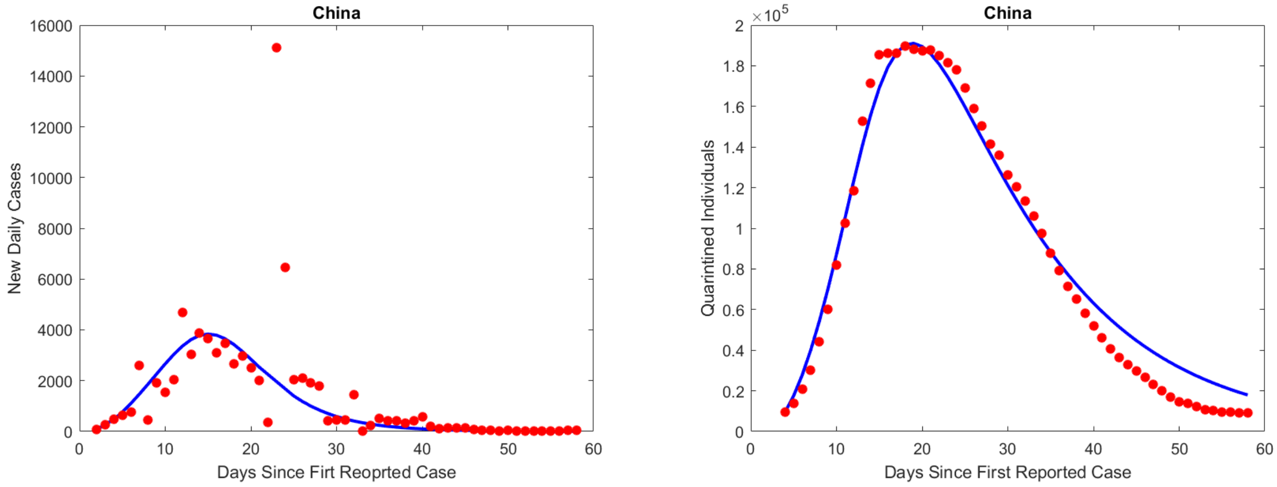


**Fig. S15.** Model fit under the assumption that $\tau$ follows an Erlang Distribution, see also table S5

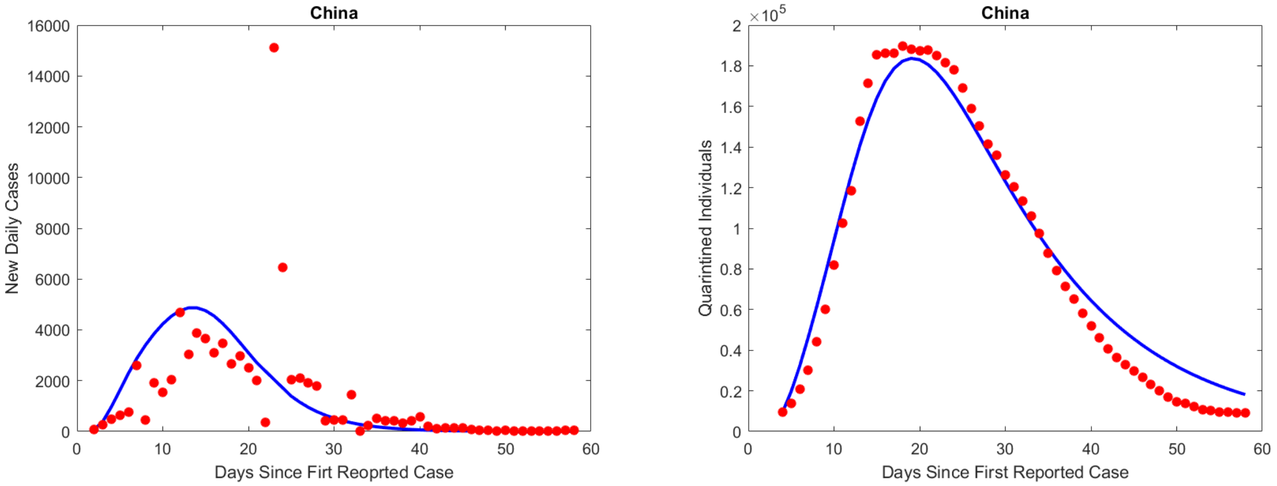


**Fig. S16.** Model fit under the assumption that $\tau$, $T$, $T_e$ all follow an Erlang Distribution, see also table S5

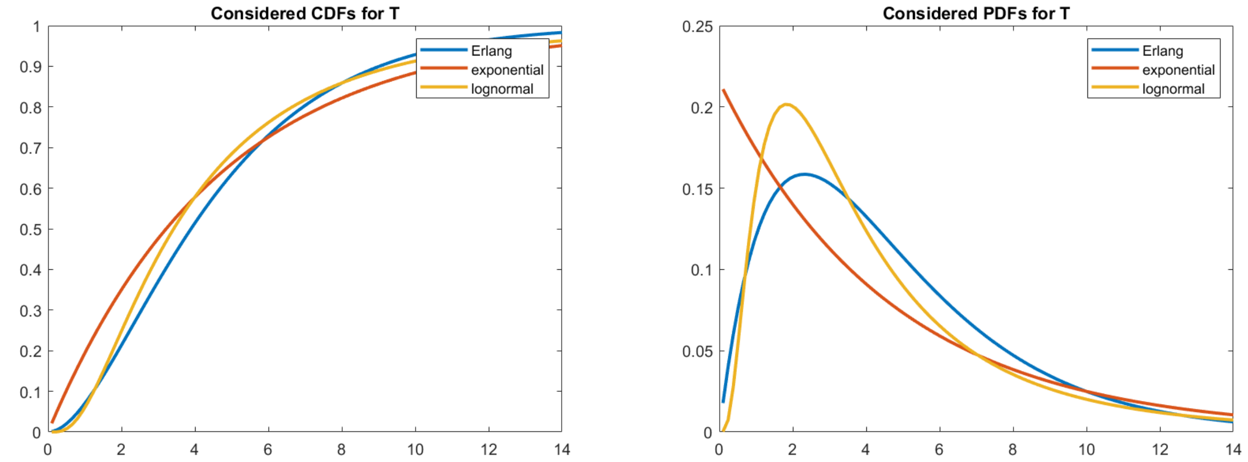


**Fig. S17.** Distribution functions for $T$, see also table S5

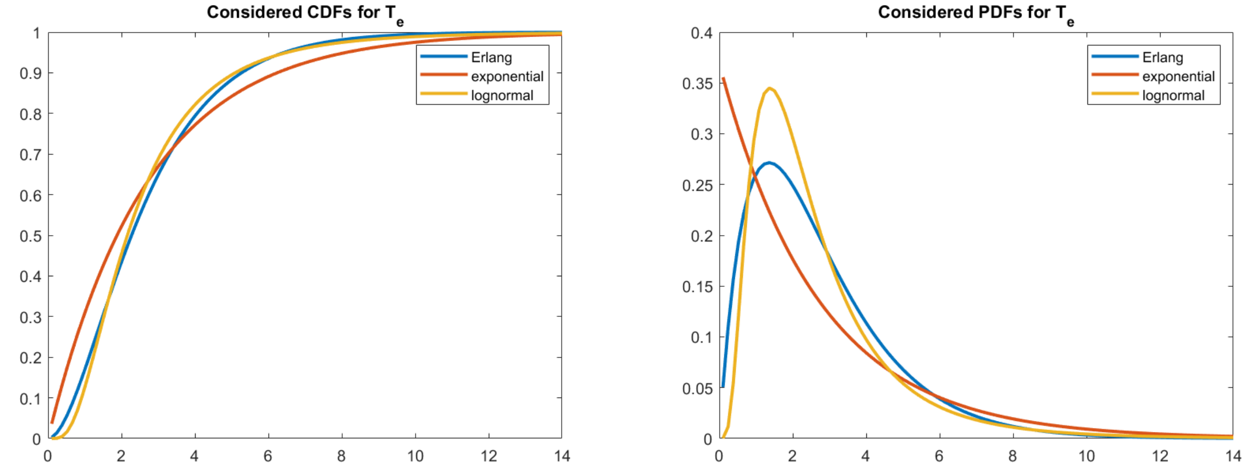


**Fig. S18.** Distribution functions for $T_e$, see also table S5

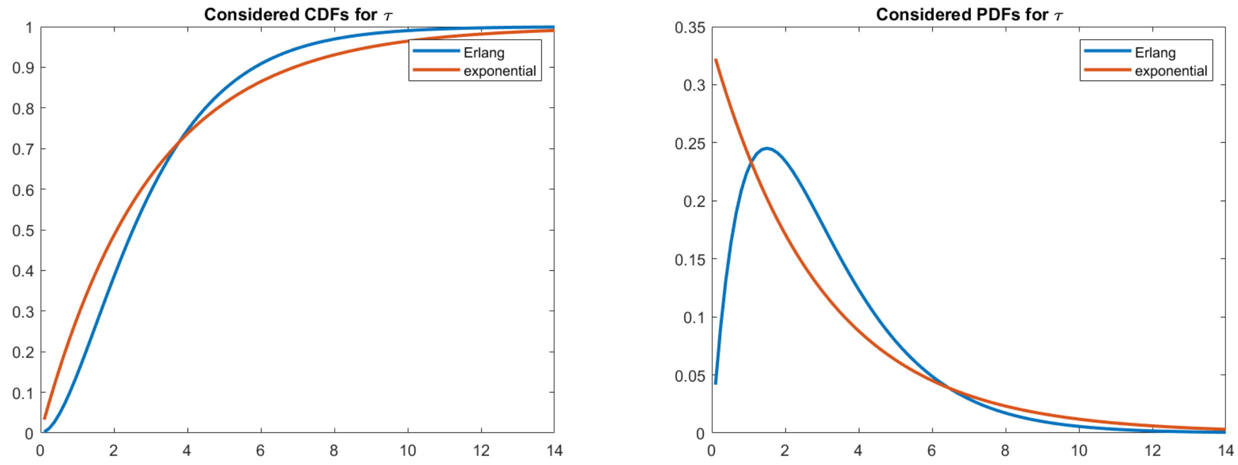


**Fig. S19.** Distribution functions for $\tau$, see also table S5

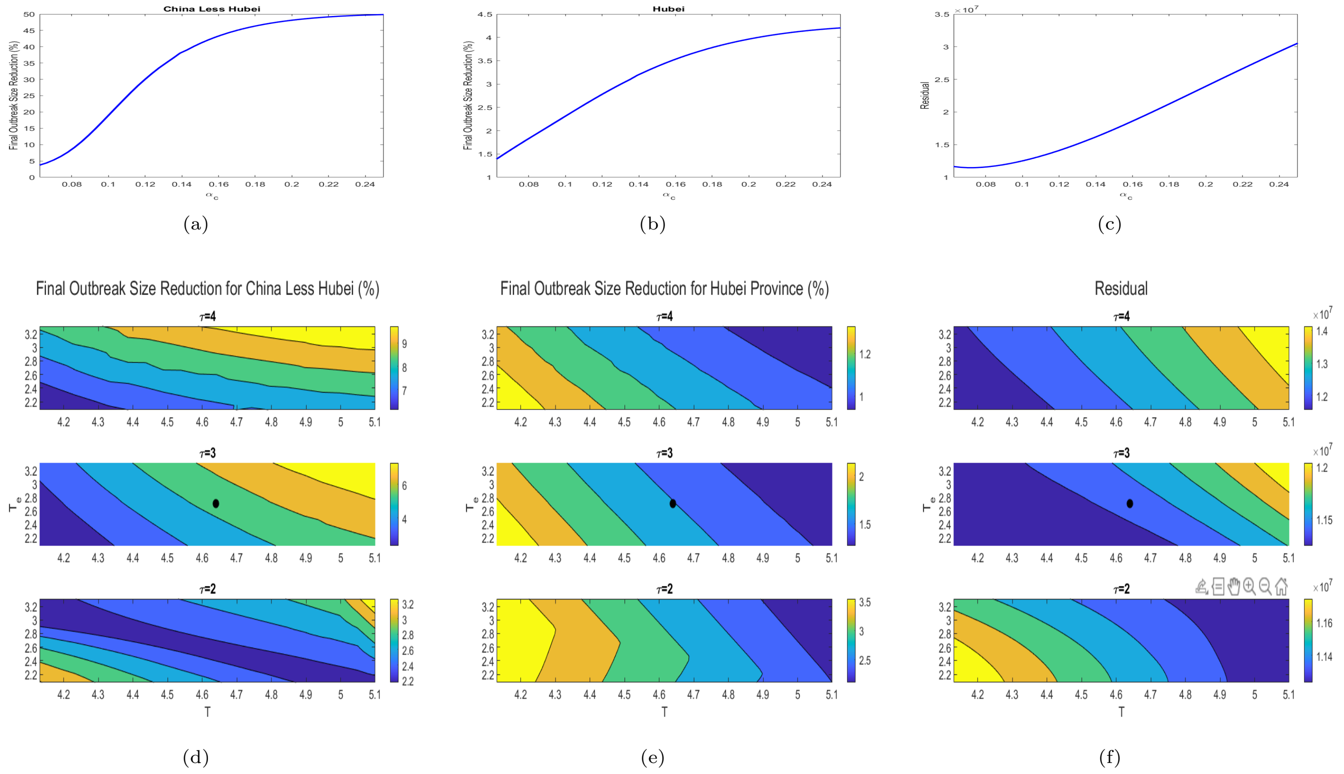


**Fig. S20.** (a-c) Examination of the effects of different $\alpha_c$ ($1/\alpha_c$ is mean quarantined contact residence time) values on model fit of Final Size Reduction **due to contact tracing** for (a) China Less Hubei and (b) Hubei, along with (c) total residual of fit. (d-f) Examination of the effects of different $T$, $T_c$, $\tau$ (infectious and exposed periods) values on model fit of Final Size Reduction **due to contact tracing** for (d) China Less Hubei and (e) Hubei, along with (f) total residual of fit.

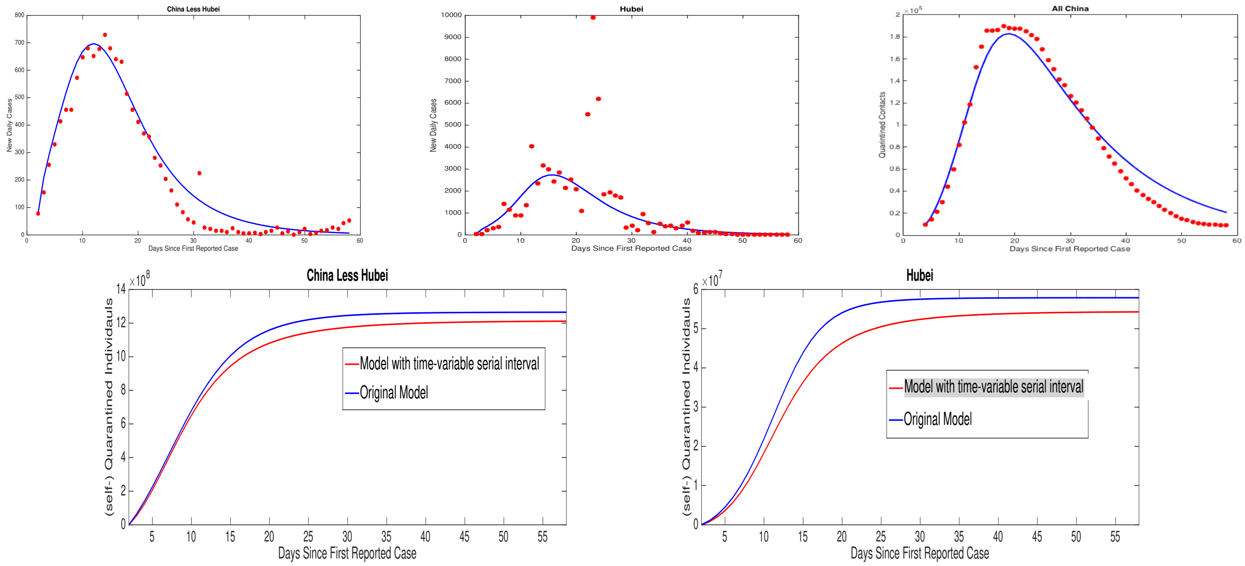


**Fig. S21.** Model fit with time variable serial interval and comparison of total self-quarantined trajectory with original fitted model with constant serial interval, see also table S5

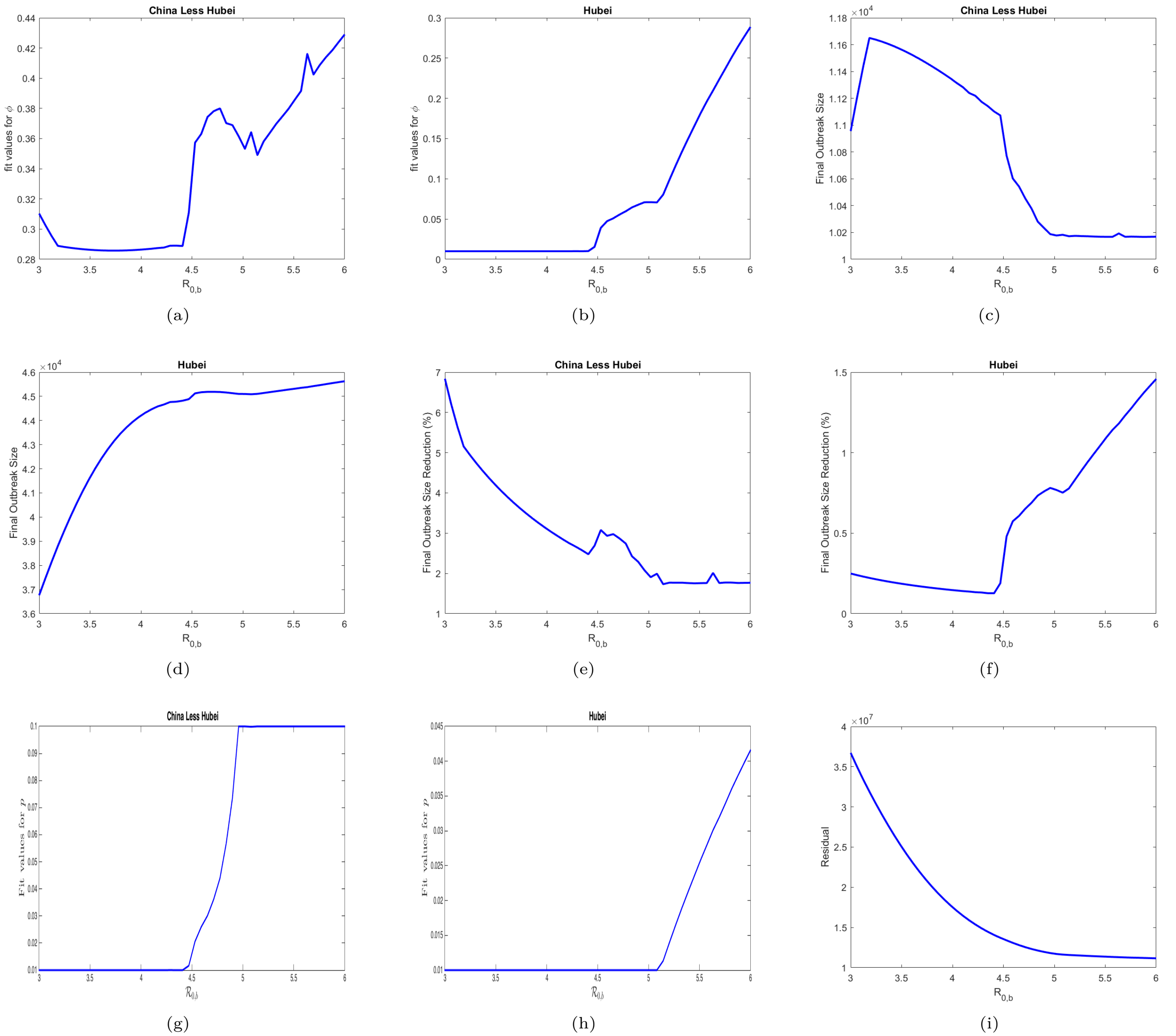


**Fig. S22.** (a) Fitted contact tracing proportion $\phi$ as a function of fixed baseline reproduction number $\mathcal{R}_{0,b}$, China Less Hubei (b)Fitted contact tracing proportion $\phi$ as a function of fixed baseline reproduction number $\mathcal{R}_{0,b}$, Hubei (c) Final Outbreak Size as a function of $\mathcal{R}_{0,b}$, China Less Hubei (d) Final Outbreak Size as a function of $\mathcal{R}_{0,b}$, Hubei (e) Reduction in Final Outbreak Size due to Contact Tracing as a function of $\mathcal{R}_{0,b}$, China Less Hubei (f) Reduction in Final Outbreak Size due to Contact Tracing as a function of $\mathcal{R}_{0,b}$, Hubei (g) $p$ vs. $\mathcal{R}_{0,b}$, China Less Hubei, (h) $p$ vs. $\mathcal{R}_{0,b}$, Hubei, (j) Residual from fitting as a function of $\mathcal{R}_{0,b}$

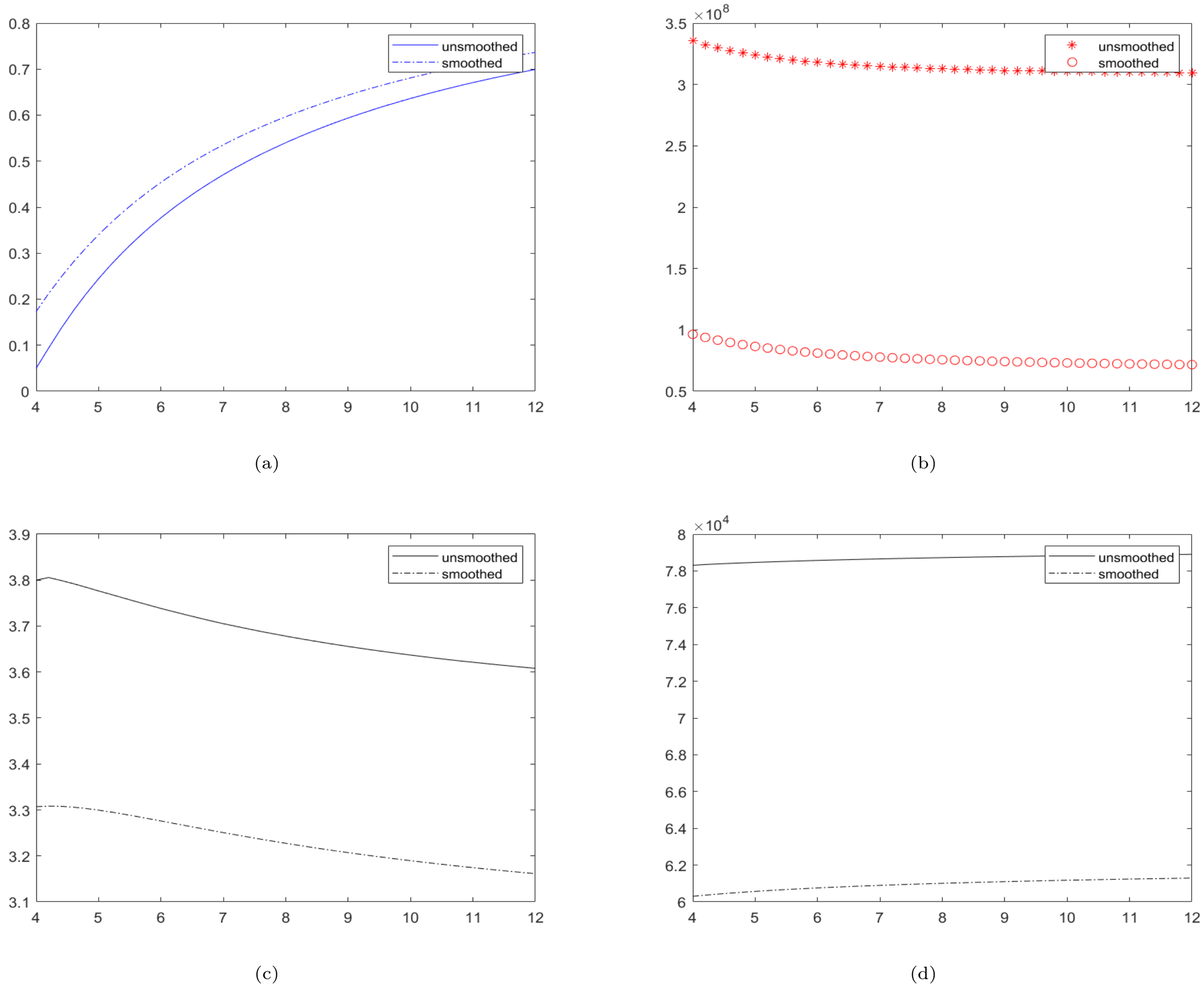


**Fig. S23.** (a) Fitted contact tracing proportion $\phi$ as a function of fixed baseline reproduction number $\mathcal{R}_{0,b}$ (b) Sum of square error from fit as a function of fixed baseline reproduction number $\mathcal{R}_{0,b}$ (c) Fitted $\mathcal{R}_0$ as a function of fixed baseline reproduction number $\mathcal{R}_{0,b}$ (d) Fitted final outbreak size as a function of fixed baseline reproduction number $\mathcal{R}_{0,b}$

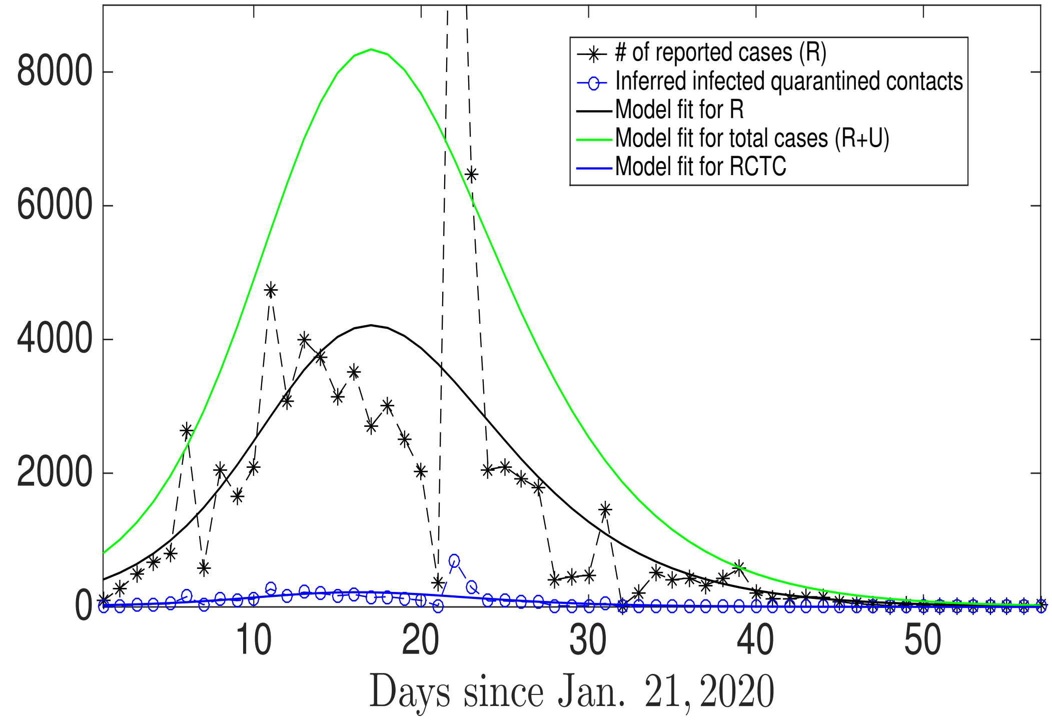


(a)

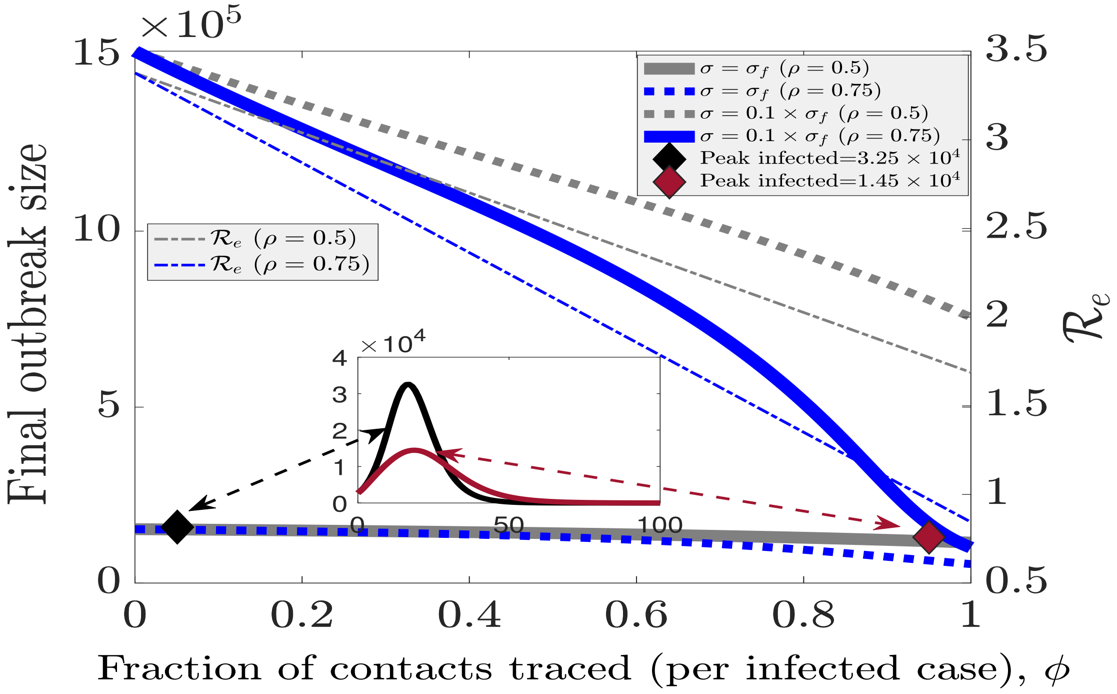


(b)

**Fig. S24.** (a) The corresponding daily reported cases and total (reported and unreported) cases in model with data and inferred subset of contact-traced infected for a fit of model incorporating unreported cases. (b) Contact tracing (CT) proportion $\phi$ versus outbreak size $\mathcal{C}_\infty$ (nonlinear relationship) and reproduction number $\mathcal{R}_0$ (linear relationship) for 2 levels of self-quarantine (SQ) rate $\sigma$ and 2 levels of reporting probability $\rho$. Note the net contact tracing probability is the product $\rho\phi$.

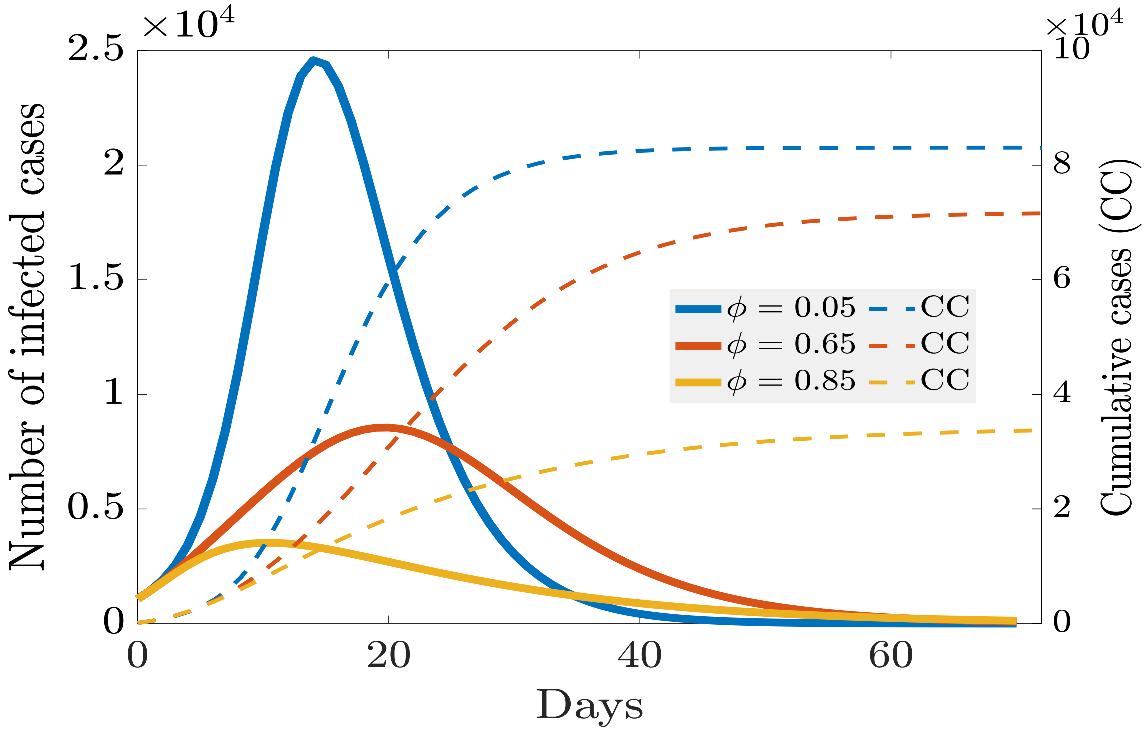


**Fig. S25.** Example epidemic curve trajectories for 3 levels of CT proportion $\phi$, corresponding to Fig.4(e)-(f) in main text, showing how contact tracing flattens the curve.

1. P Van den Driessche, J Watmough, Reproduction numbers and sub-threshold endemic equilibria for compartmental models of disease transmission. *Mathematical biosciences* **180**, 29–48 (2002).
2. J Arino, F Brauer, P Van Den Driessche, J Watmough, J Wu, A final size relation for epidemic models. *Mathematical Biosciences & Engineering* **4**, 159 (2007).
3. Z Feng, Final and peak epidemic sizes for seir models with quarantine and isolation. *Mathematical Biosciences & Engineering* **4**, 675 (2007).
4. NHC, National Health Commission of the People's Republic of China (http://en.nhc.gov.cn) (2020).
5. TK Tsang, et al., Effect of changing case definitions for covid-19 on the epidemic curve and transmission parameters in mainland china: a modelling study. *The Lancet Public Health* (2020).
6. AA King, M Domenech de Cellès, FM Magpantay, P Rohani, Avoidable errors in the modelling of outbreaks of emerging pathogens, with special reference to ebola. *Proceedings of the Royal Society B: Biological Sciences* **282**, 20150347 (2015).
7. Q Bi, et al., Epidemiology and transmission of covid-19 in 391 cases and 1286 of their close contacts in shenzhen, china: a retrospective cohort study. *The Lancet Infectious Diseases* (2020).
8. K Sun, et al., Transmission heterogeneities, kinetics, and controllability of sars-cov-2. *medRxiv* (2020).
9. C Browne, H Gulbudak, G Webb, Modeling contact tracing in outbreaks with application to ebola. *Journal of theoretical biology* **384**, 33–49 (2015).
10. J Wallinga, P Teunis, Different epidemic curves for severe acute respiratory syndrome reveal similar impacts of control measures. *American Journal of epidemiology* **160**, 509–516 (2004).
11. CD Lab, Baidu Mobility Data (2020).
12. L Luo, et al., Contact settings and risk for transmission in 3410 close contacts of patients with covid-19 in guangzhou, china: A prospective cohort study. *Annals of internal medicine* (year?).
13. ST Ali, et al., Serial interval of sars-cov-2 was shortened over time by nonpharmaceutical interventions. *Science* **369**, 1106–1109 (2020).
14. X He, et al., Temporal dynamics in viral shedding and transmissibility of covid-19. *Nature medicine* **26**, 672–675 (2020).
15. BF Maier, D Brockmann, Effective containment explains subexponential growth in recent confirmed covid-19 cases in china. *Science* **368**, 742–746 (2020).
16. NB of Statistics of China, Provincial Population Data (2019).
